# Rare variants in *PPFIA3* cause delayed development, intellectual disability, autism, and epilepsy

**DOI:** 10.1101/2023.03.27.23287689

**Authors:** Maimuna S. Paul, Sydney L. Michener, Hongling Pan, Jessica M. Pfliger, Jill A. Rosenfeld, Vanesa C. Lerma, Alyssa Tran, Megan A. Longley, Richard A. Lewis, Monika Weisz-Hubshman, Mir Reza Bekheirnia, Nasim Bekheirnia, Lauren Massingham, Michael Zech, Matias Wagner, Hartmut Engels, Kirsten Cremer, Elisabeth Mangold, Sophia Peters, Jessica Trautmann, Jessica L. Mester, Maria J. Guillen Sacoto, Richard Person, Pamela P. McDonnell, Stacey R. Cohen, Laina Lusk, Ana S.A. Cohen, Jean-Baptiste Le Pichon, Tomi Pastinen, Dihong Zhou, Kendra Engleman, Caroline Racine, Laurence Faivre, Sébastien Moutton, Anne-Sophie Denommé- Pichon, Sarah Schuhmann, Georgia Vasileiou, Sophie Russ-Hall, Ingrid E. Scheffer, Gemma L. Carvill, Heather Mefford, Undiagnosed Diseases Network, Carlos A. Bacino, Brendan H. Lee, Hsiao-Tuan Chao

## Abstract

*PPFIA3* encodes the Protein-Tyrosine Phosphatase, Receptor-Type, F Polypeptide-Interacting Protein Alpha-3 (PPFIA3), which is a member of the LAR protein-tyrosine phosphatase-interacting protein (liprin) family involved in synaptic vesicle transport and presynaptic active zone assembly. The protein structure and function are well conserved in both invertebrates and vertebrates, but human diseases related to PPFIA3 dysfunction are not yet known. Here, we report 14 individuals with rare mono-allelic *PPFIA3* variants presenting with features including developmental delay, intellectual disability, hypotonia, autism, and epilepsy. To determine the pathogenicity of *PPFIA3* variants *in vivo*, we generated transgenic fruit flies expressing either human PPFIA3 wildtype (WT) or variant protein using GAL4-UAS targeted gene expression systems. Ubiquitous expression with Actin-GAL4 showed that the *PPFIA3* variants had variable penetrance of pupal lethality, eclosion defects, and anatomical leg defects. Neuronal expression with elav-GAL4 showed that the *PPFIA3* variants had seizure-like behaviors, motor defects, and bouton loss at the 3^rd^ instar larval neuromuscular junction (NMJ). Altogether, in the fly overexpression assays, we found that the *PPFIA3* variants in the N-terminal coiled coil domain exhibited stronger phenotypes compared to those in the C-terminal region. In the loss-of-function fly assay, we show that the homozygous loss of fly *Liprin-*α leads to embryonic lethality. This lethality is partially rescued by the expression of human *PPFIA3* WT, suggesting human PPFIA3 protein function is partially conserved in the fly. However, the *PPFIA3* variants failed to rescue lethality. Altogether, the human and fruit fly data reveal that the rare *PPFIA3* variants are dominant negative loss-of-function alleles that perturb multiple developmental processes and synapse formation.

## Introduction

Synapses are highly specialized communication junctions between neurons and their target cells where neurotransmitter release occurs in an intricately coordinated manner. In the presynaptic neuron, a key site for neurotransmitter release is the active zone, which is composed of a complex protein matrix.^1–4^ RIM, ELKS, Munc13, RIM-BP, Piccolo/Bassoon, and Liprin-α are the six major protein families comprising the active zone.^5^ These active zone proteins along with other cytoskeletal proteins, Ca^2+^ channels, and SNAREs (soluble N-ethylmaleimide-sensitive fusion protein attachment protein receptors) form a tightly orchestrated unit to mediate synaptic vesicle docking, priming, fusion, and neurotransmitter release.^5^ Disruption of synapse structure or function leading to variable defects in neurotransmitter release is widely responsible for neurodevelopmental and neuropsychiatric disorders including epilepsy, intellectual disability (ID), autism spectrum disorder (ASD), schizophrenia, and bipolar disorder.^6–10^

The network of multidomain proteins comprising the active zone falls into different categories such as cytoskeletal and scaffolding proteins, adhesion molecules, calcium channels, and synaptic vesicle release machinery. Liprins are a critical class of scaffolding proteins found in the active zone. In conjunction with the adhesion molecule LAR-PTPs (Leukocyte Antigen Receptor-Protein Tyrosine Phosphatases), liprins play a key role in the active zone organization and structure. Liprin family members are identified as interactors of LAR-PTPs and are subdivided into liprin-α and liprin-β proteins.^11, 12^ Structural studies show that liprins are comprised of an N-terminal coiled coil domain and C-terminal sterile-α-motif (SAM) domain.^11, 12^ The N-terminal coiled coil domain mediates homodimerization and heterodimerization with other liprin-α members and interactions with other active zone proteins such as RIM and ELKS.^13–16^ In contrast, the C-terminal SAM domain of liprin-α interacts with the LAR intracellular domain.^17^ Additionally, liprins interact with kinesin motor proteins^18–20^ and are involved in the hedgehog signaling-dependent trafficking of Kif7 and Gli to the cilia in the context of embryonic development and cortical microtubule organization.^18, 19^

Liprins are also known as protein-tyrosine phosphatase, receptor-type, f polypeptide (PTPRF)-interacting protein α (PPFIA) or β (PPFIB). Vertebrates have four Ppfia (1-4) and two Ppfib (1-2) proteins that are encoded by *Ppfia1-4* or *Ppfib1-2* respectively.^12^ Expression studies in mice show that all four mouse Ppfia1-4 homologs are expressed in the brain, with differences in distribution and expression levels.^21^ Ppfia1 is expressed in the brain, lung, heart, liver, muscle, spleen, and testes.^12, 22, 23^ Ppfia2, Ppfia3, and Ppfia4 are predominantly expressed in the brain,^12, 22, 23^ including structures such as the olfactory bulb, striatum, cortex, hippocampus, thalamus, midbrain, cerebellum, and brainstem.^21–23^ In contrast, Ppfia1 expression in the brain is predominantly localized to the cerebellum and olfactory bulb.^21–23^ A subcellular localization study showed that Ppfia2 and Ppfia3 are located in both the pre-synaptic and post-synaptic compartments.^23^ However, only Ppfia3 specifically colocalizes in the presynaptic compartment and mediates protein-protein interactions with the active zone proteins Bassoon, RIM, Munc-13, RIM-BP, and ELKS in hippocampal neurons.^24^

Ppfia proteins are well conserved in both vertebrates and invertebrates. Studies in *C. elegans* identified that the sole Ppfia homolog, syd-2, plays a key role in presynaptic active zone organization.^25, 26^ Studies showed that syd-2 recruits synaptic components to presynaptic sites and contributes to the formation of neuromuscular junctions (NMJs), along with active zone assembly and stabilization.^26, 27^ Mutant syd-2 worms show presynaptic active zone defects due to disruption of syd-2 oligomerization.^13, 26, 27^ A similar role was found for the fruit fly homolog, Liprin-α, where it is required for synapse formation, synaptic vesicular transport, active zone assembly, and axonal target selection in the retina.^28–30^ Consistent with the invertebrate models, synaptic ultrastructure and electrophysiological studies in *Ppfia3* knock-out mice found impaired presynaptic active zone assembly and synaptic vesicle docking, tethering, and exocytosis.^24^ Altogether, these studies reveal that Ppfia family members are integral scaffolding proteins for the assembly of intricate protein complexes involved in synapse formation, synaptic transmission, and protein trafficking.

Here, we report a cohort of 14 individuals from 12 families with rare mono-allelic variants in *PPFIA3* associated with developmental delay (DD) (11/14), ID (9/14), dysmorphisms (8/14), hypotonia (6/14), ASD (6/14), and abnormal electroencephalogram (EEG) or epilepsy (5/14). In these 14 individuals, 11 have *PPFIA3* missense variants where ten variants are *de novo* and one with unknown inheritance, one individual has a frameshift variant with unknown inheritance, and two related individuals have a heterozygous splice variant. The phenotypic consequences of rare variants in *PPFIA3* are currently unknown. Molecular modeling of *PPFIA3* missense variants showed that the amino acid changes disrupt the polar contacts and residue interactions suggesting these alterations would be deleterious to PPFIA3 protein function. To determine the pathogenicity of the *PPFIA3* variants *in vivo,* we used fruit fly functional assays where we examined five of the *de novo PPFIA3* missense variants, GenBank NM_003660.4: c.115C>T [p.(Arg39Cys)], c.943G>T [p.(Ala315Ser)], c.1243C>T [p.(Arg415Trp)], c.1638G>T [p.(Trp546Cys)], and c.2350C>T [p.(Arg784Trp)]. Overexpression fly assays revealed that the *PPFIA3* variants are associated with behavioral, developmental, and NMJ defects. Loss-of-function (LOF) fly assays recapitulated an embryonic lethality phenotype induced by loss of fly *Liprin-*α, which was partially rescued with human *PPFIA3* wildtype (WT) cDNA expression. In contrast, expression of the *PPFIA3* variant cDNAs either failed to rescue or partially rescued the embryonic lethality compared to *PPFIA3* WT cDNA. Altogether, we show that rare *PPFIA3* variants are deleterious to protein function with *in vivo* fruit fly assays and lead to an autosomal dominant neurodevelopmental disorder characterized by DD/ID, ASD, and epilepsy in humans.

## Materials and Methods

### Human subjects

Clinical data were acquired after written informed consent to participate in the study and publication of results was obtained from the participant or their legal representative in accordance with the ethical standards of the participating institutional review boards (IRB) on human research at each respective institution. GeneMatcher was used to form an international collaboration, allowing for comparison of individuals and their variants.^31–33^ Collection and analysis of the de-identified clinical cohort was approved by Baylor College of Medicine’s IRB. *PPFIA3* heterozygous variants were identified by exome sequencing (ES) through each individual’s respective institution. DNA was extracted from peripheral blood mononuclear cells or buccal sample for ES. Exome or Sanger sequencing of the parental samples were performed when feasible to confirm *de novo* or inherited segregation. Paternity was confirmed by the inheritance of rare single nucleotide polymorphisms from the parents. Sample swap was excluded. Participant ID’s are not known to anyone outside of the research group.

### Molecular modeling

Molecular visualization of the PPFIA3 protein structure was completed with PyMol (The PyMOL Molecular Graphics System, Version 2.5.2 Schrödinger, LLC.). The crystal structure of PPFIA3 (GenBank NP_003651.1, Uniprot ID: O75145) was used to build the PPFIA3 protein structure model in PyMol. Affected residues were altered to the corresponding human variants and the mutation effects were modeled alongside the native protein. The changes in the PPFIA3 protein structure were assessed by displaying local polar contacts and residue interactions before and after mutagenesis.

### *Drosophila melanogaster* stocks and maintenance

All the fruit fly stocks used in this study were reared in standard cornmeal and molasses-based fly food at room temperature (RT, 20-21°C) unless otherwise noted. The fruit fly stocks used in the study were either obtained from Bloomington Drosophila Stock Center (BDSC) or generated at the Jan and Dan Duncan Neurological Research Institute. We generated transgenic fly alleles as previously described^34^ by utilizing the pUASg-HA-attB vector^35^ to express the human *PPFIA3* WT and variant cDNAs with a C-terminal hemagglutinin (HA) tag under the control of upstream activating system (UAS) elements by Gateway LR Cloning (LR Clonase II, Thermo Fisher Scientific, Cat #11791020). To generate the *PPFIA3* variants, we utilized the human full-length cDNA of *PPFIA3* (GenBank: NM_003660.4). *PPFIA3* c.115 C>T [p.(Arg39Cys)], *PPFIA3* c.943G>T [p.(Ala315Ser)], *PPFIA3* c.1243C>T [p.(Arg415Trp)], *PPFIA3* c.1638G>T [p.(Trp546Cys)], and *PPFIA3* c.2350C>T [p.(Arg784Trp)] were generated by Q5 site-directed mutagenesis (New England Biolabs, Cat #M0491S) in the pDONR221 Gateway compatible donor vector. The constructs were confirmed by Sanger sequencing. Primer sequences for the site-directed mutagenesis and Sanger sequencing are listed in **Table S1**. Human *PPFIA3* WT and variant cDNAs were inserted into the chromosome-3 VK33 (PBac{y[+]-attP}VK00033) docking site by φC31-mediated recombination for fruit fly transgenesis.^35^ Transgenic UAS fly alleles generated in this study include *UAS-PPFIA3-WT-HA, UAS-PPFIA3-p.(Arg39Cys)-HA, UAS-PPFIA3-p.(Ala315Ser)-HA, UAS-PPFIA3-p.(Arg415Trp)-HA,* and *UAS-PPFIA3-p.(Trp546Cys)-HA,* and *UAS-PPFIA3-p.(Arg784Trp)-HA*. Fly alleles from the stock centers include: *Liprin-*α*^F3ex15^/In(2LR)Gla* (BDSC#8563), *w[1118]; Df(2L)Exel7027/CyO* (BDSC#7801), *y[1] w[118]; PBac{y[+]-aatP-3B}-VK00033* (BDSC#9750), and *elav-GAL4/CyO* (BDSC#8765). *UAS-empty-VK33, Actin-GAL4,* and *da-GAL4* lines were obtained from Dr. Hugo J. Bellen.

### Larval brain and NMJ immunostaining and confocal microscopy

Fruit fly larval brains or whole-body wall muscles including the central nervous system were dissected from wandering third instar larvae reared at 25°C in ice-cold 1X-PBS and fixed in 4%-paraformaldehyde for 20 minutes at RT. The tissues were washed four times in Tri-PBS (1X-PBS + 0.2% Triton-X-100) with 1%-Bovine Serum Albumin (BSA) for 15-minutes each followed by incubation in blocking solution (Tri-PBS with 0.1% BSA and 8% normal donkey serum) for 30 minutes. Primary antibodies, rat anti-HA (1:50, clone 3F10, Millipore Sigma, Cat#11867423001), mouse anti-elav (1:100, Developmental Studies Hybridoma Bank, Cat#9F8A9), mouse anti-Bruchpilot (Brp) (1:50, Developmental Studies Hybridoma Bank, Cat#nc82), and goat anti-Horseradish Peroxidase (HRP) (1:1000, Jackson ImmunoResearch, Cat#123-005-021) were diluted in blocking solution, added to the tissues, and incubated overnight at 4°C. The tissues were rinsed three to four times in Tri-PBS with 1%-BSA for 15 minutes each followed by incubation in blocking solution for 30 minutes at RT. The secondary antibodies, donkey anti-rat IgG antibody (Cy3) (1:300, Jackson ImmunoResearch, Cat#712-165-153), Alexa Fluor 488 Affinipure donkey anti-goat IgG (H+L) (1:300, Jackson ImmunoResearch, Cat#705-545-147) and Alexa Fluor 488 Affinipure donkey anti-mouse IgG (H+L) (1:300, Jackson ImmunoResearch, Cat#715-545-151) were diluted in blocking solution and added to the tissues for a 90-minute incubation at RT on a rocker. For NMJ staining, phalloidin (Phalloidin-iFluor 405 Reagent, Abcam, Cat#ab176752) was added along with the secondary antibodies to visualize the muscles. After removing the secondary antibody, tissues were washed three times in Tri-PBS with 1% BSA for 15 minutes each, and then rinsed in 1X-PBS at RT. For larval brains, this was followed by incubation in 406-diamidino-2-phenylindole dihydrochloride (DAPI, 1 mg/mL, Cayman Chemical, Cat#14285) for 30 minutes at RT. After removing DAPI, a final wash was completed with 1X-PBS for 15 minutes at RT. The tissues were mounted in Prolong Glass anti-fade mountant (Thermo Scientific, Cat#36984). Images were acquired on a Leica Sp8 laser-scanning confocal microscope. The same settings for laser power and detector gain were used for all genotypes. Third instar larval brain images were acquired as a z-stack with a z-step of 1µm and line average of four at 400 Hz with a 20X objective at 1024 x 1024-pixel resolution. NMJ images were acquired with a 40X objective. Maximum intensity projections were created from the z-stack in ImageJ. All images were processed and assembled using ImageJ and Adobe Illustrator.

### 3^rd^ instar larval NMJ quantifications

The total number of boutons from abdominal segment A3, muscle 6/7 were counted manually using Imaris. The spot function was used with point style sphere and radius scale 1.0 to count the number of boutons. The NMJ length was quantified using the HRP staining and measured in ImageJ. Data was collected and analyzed blinded to genotypes. Statistical analysis between the control and experimental groups was conducted with one-way ANOVA and Tukey’s post-hoc analysis in GraphPad Prism 8.

### Adult fruit fly behavioral assays

For the climbing assay, 15-day-old flies of both sexes were anesthetized with CO_2_ 24 hours prior to being tested and two to three flies were housed in food-containing vials at room temperature. At the time of assay, these flies were transferred without anesthesia to a clear graduated cylinder with a 20-cm mark. The flies were tapped three times to the bottom of the cylinder to examine negative geotaxis (climbing upward). The cutoff time to reach the 20-cm mark was 40 seconds. A total of 40-55 flies of both sexes were tested for each genotype. Crosses for the climbing assay were set up at 25°C and the assay was performed at 20-21°C.

For the bang sensitivity assay, 15-day-old flies of both sexes were anesthetized with CO_2_ 24 hours prior to being tested and two to three flies were housed in food-containing vials at room temperature. At the time of assay, these flies were transferred without anesthesia to an empty food vial and vortexed for 15 seconds. Flies were observed for time to recover from the vortexing. The cutoff time to recover was 30 seconds. Recovery was defined as being upright and mobile. Flies were considered bang sensitive if they remained upside down, immobile, or showing rhythmic involuntary movements suggestive of seizure-like behaviors. A total of 40-55 flies of both sexes were tested for each genotype. Crosses for the bang sensitivity assay were set up at 25°C and the assay was performed at 20-21°C.

### Adult fruit fly leg mounting

Adult flies were fixed overnight in ethanol at room temperature and the legs were dissected and mounted using CMCP-10 Macroinvertebrate High Viscosity Mountant (D/S259) (Electron Microscopy Sciences, Cat#18004-02). Leg images were taken using the Leica MZ16 stereomicroscope. Images were processed and assembled using Adobe Photoshop CS5.1 and Adobe illustrator. Crosses were set up at 25°C.

### Genomic DNA isolation and PCR

Genomic DNA was extracted by homogenizing three whole flies in 50mM sodium hydroxide and heating the samples at 95°C for 30 minutes followed by the addition of 1M Tris-HCl (pH 7.5) to stop the lysis. 100 ng of genomic DNA was used to amplify the *HA*-, *PPFIA3*-, and *rps17*-specific sequences. The experiment was repeated in three independent biological replicates. *HA* and *PPFIA3* band intensity were quantified by normalizing to the band intensity of the endogenous reference gene *rps17* and plotted as fold-change relative to the control. Flies were maintained at 20-21°C. Primer sequences are listed in **Table S1**.

## Results

### Identification of rare mono-allelic *PPFIA3* variants in individuals with developmental delay, intellectual disability, hypotonia, autism, and epilepsy

An international collaboration through the Undiagnosed Diseases Network (UDN)^36^ and GeneMatcher^31–33^ led to the identification of 14 individuals with neurodevelopmental phenotypes and rare missense, frameshift deletion, or consensus splice mono-allelic variants in *PPFIA3* (**Figure 1A, B**). The variants from all affected individuals were identified through exome sequencing (ES) or Sanger sequencing. Ten of the individuals harbored *de novo* (I:1, and I:5-13) missense variants, one individual (I:3) inherited a consensus splice variant from a symptomatic parent (I:4), and the inheritance pattern for three affected individuals is unknown (I:2, I:4, and I:14) (**Table 1**). The individuals with unknown inheritance pattern have either a missense variant (I:2), a consensus splice variant (I:4), or a frameshift deletion (I:14). The *de novo* p.(Arg429Trp) variant was seen in a mosaic state (present in 26% of the reads from ES, suggesting heterozygosity in ∼52% of cells) in I:7. The Combined Annotation Depletion Score (CADD)^37^ for the variants ranged from 21.1 to 35 (**Table S2**). Thirteen out of fourteen variants were absent from the Genome Aggregation Database (gnomAD, v2.1.1).^38^ The p.(Arg784Trp) variant had a frequency of 3.19×10^-5^ (1/31,386) in gnomAD (v2.1.1) and was identified as a *de novo* finding in an individual (I:10) with mild ID and Landau-Kleffner epilepsy syndrome. We also identified a p.(Arg559Trp) variant in an individual (I:15) with a discordant severe neurodegenerative phenotype and unknown inheritance of the variant (**Table S3**). The p.(Arg559Trp) variant was absent from gnomAD (v2.1.1).

**Figure 1:**
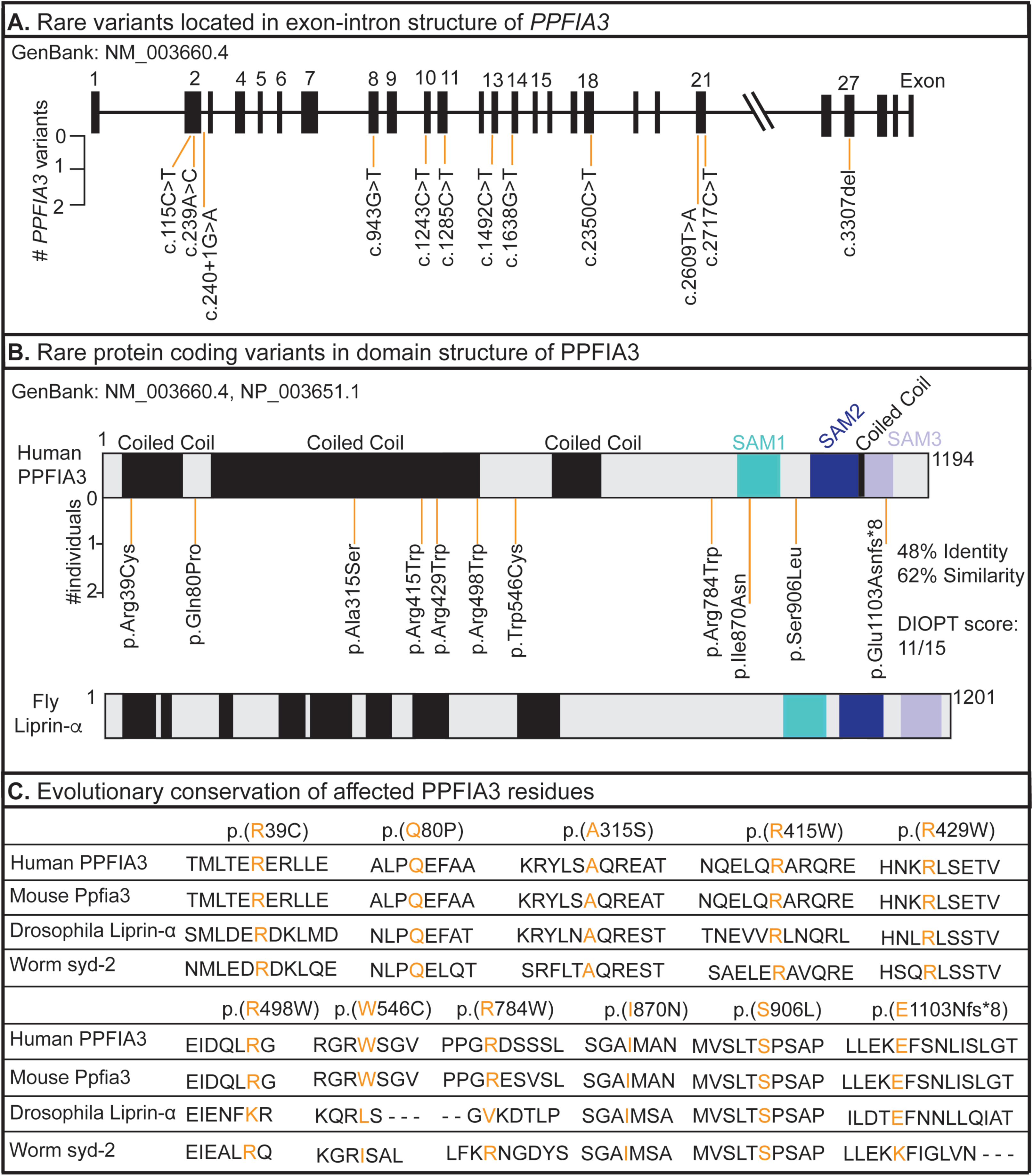
Variant location and evolutionary conservation of the affected protein residue between human PPFIA3 and orthologs in vertebrate and invertebrate species. (A) Location of *PPFIA3* variants in the genomic locus corresponding to the exon-intron structure. Number of individuals with the rare mono-allelic *PPFIA3* variant shown in the y-axis. **(B)** Location of *PPFIA3* variants in the corresponding protein domains. Number of individuals with the variant shown in the y-axis. The fruit fly homolog, *Liprin-*α, shows 48% identity and 62% similarity with the human *PPFIA3*. Sterile alpha motif (SAM). **(C)** Conservation analysis of affected residues from the 14 individuals with neurodevelopmental findings are shown in *H. sapiens* (human), *M. musculus* (mouse), *D. melanogaster* (Drosophila), and *C. elegans* (worm) with the affected residue in orange. Residues within or adjacent to a functional domain (coiled coil or SAM domains) are highly conserved across vertebrates and invertebrates.

**Table 1:**
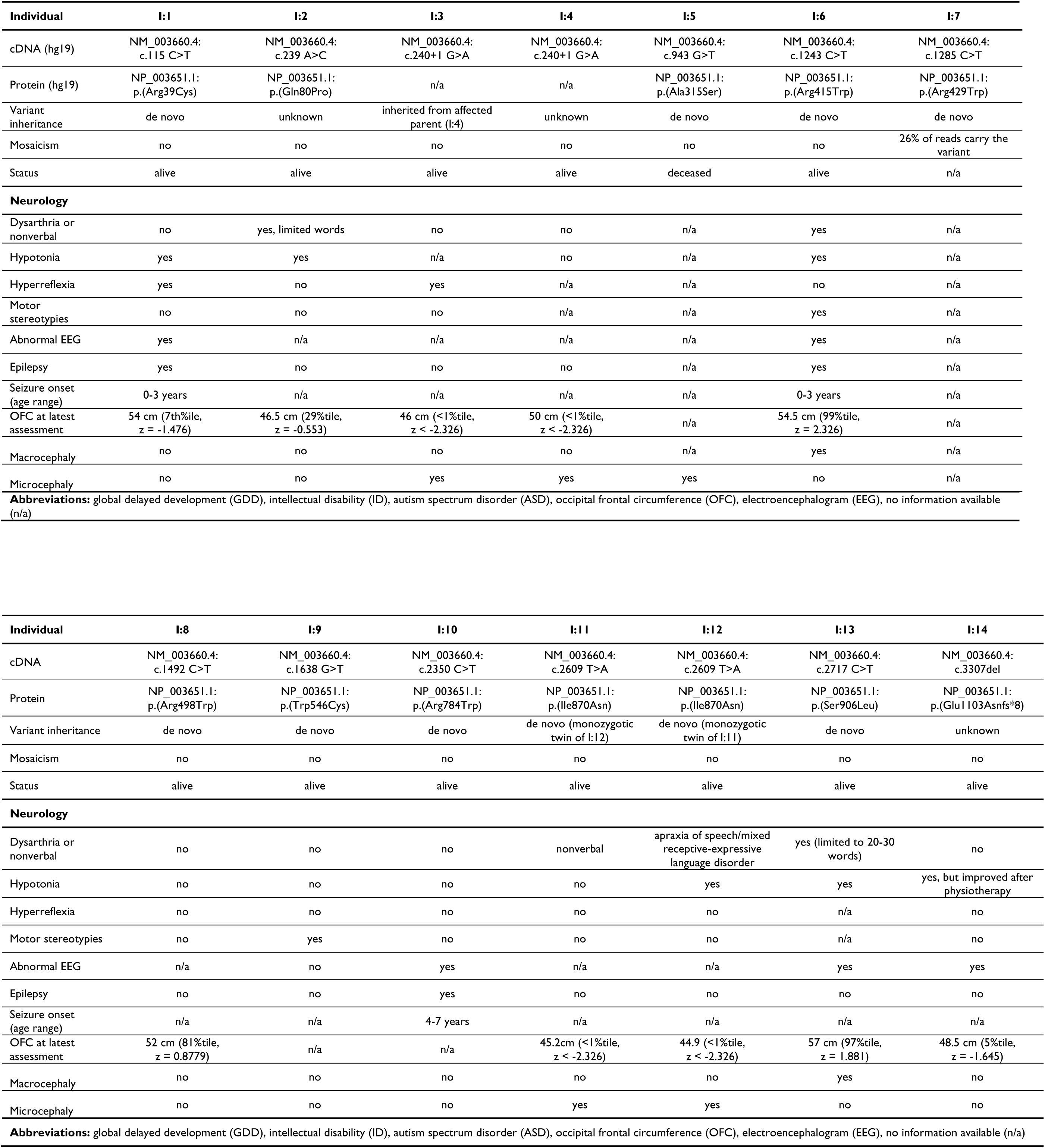
Genetic and neurologic findings in individuals with rare mono-allelic *PPFIA3* variants and neurodevelopmental phenotypes. . Molecular findings, demographics, and neurology findings from individuals I:1 - I:14 with *PPFIA3* variants.

In a statistical model of *de novo* variants for autism spectrum and intellectual disability disorders (ASD/ID), the model identified *PPFIA3* as one of ∼1,000 genes significantly lacking functional variation in non-ASD/ID individuals but are enriched with *de novo* variants in ASD/ID individuals.^39^ Furthermore, gnomAD v2.1.1. analysis showed that *PPFIA3* has a high probability of loss-of-function (LOF) intolerance (LOEUF = 0.12, pLI = 1.0), as 64.1 LOF variants were expected given the gene size and GC content but only three LOF variants were observed.^38^ *PPFIA3* is also a highly constrained gene with a missense z-score of 5.49, suggesting intolerance to missense variation, as 727.5 missense variants were expected but only 311 were observed.^38^ Together, these findings support that the rare mono-allelic *PPFIA3* variants may cause a neurodevelopmental phenotype.

Twelve individuals in the cohort had DD/ID (I:1-I:4, I:6, I:8-14) (**Tables S4, S5**), while the remaining two individuals (I:5 and I:7) could not be assessed for this feature due to early death. Individual I:5 had renal failure, severe anorectal malformation with complete anal atresia, absent bladder, and dysmorphisms, and death in infancy. Individual I:7 had a prenatal diagnosis of abnormal gyration and ventriculomegaly, which led to elective pregnancy termination. Abnormal EEG (5/14; I:1, I:6, I:10, I:13-14) and epilepsy (3/14; I:1, I:6, I:10) findings were common (**Table 1**). The affected individuals had multiple seizure semiologies including focal clonic seizures, atonic seizures, absence seizures, and focal tonic-clonic seizures with secondary generalization (**Tables 1, S4, S5**). Three probands had neuroanatomical changes detected by MRI (I:6-7, I:12), which included flattening of the posterior globes at the level of optic nerve insertion, abnormal gyration with ventriculomegaly, and mild periventricular leukomalacia with mild white matter volume loss (**Table S6)**. Delayed speech development was present in ten individuals (I:1-3, I:6, I:8, I:10-14) with absent expressive language in one individual (I:6) (**Tables S4**, **S5**). Hypotonia was present in six individuals (I:1-2, I:6, I:12-14) (**Table 1**). Co-morbid ASD diagnosis was reported in six individuals (I:2, I:6, I:8-9, I:11, I:13) (**Tables S4, S5**). Gastrointestinal dysmotility characterized by constipation, difficulty feeding, and dysphagia was present in seven individuals (I:1-2, I:5-6, I:10-12) (**Tables S4, S5**). Dysmorphic facial features were present in six individuals, and included prominent forehead, strabismus, and bilateral epicanthal folds (I:1-3, I:6, I:8 I:14) (**Tables S4, S5**). Macrocephaly or microcephaly were present in seven individuals (I:3-6, I:11-13) (**Tables 1, S4, S5**).

### Conservation analysis and molecular modeling of *PPFIA3* variants

The predicted pathogenicity of *PPFIA3* variants was validated *in vivo* using *D. melanogaster* (fruit fly). The fly homolog of *PPFIA3* is *Liprin-*α, and the fly protein shows an overall 48% identity and 62% similarity with the human protein (**Figure 1B**). Similar to the human PPFIA3 protein, the fruit fly Liprin-α contains N-terminal coiled coil domains and three C-terminal SAM domains (**Figure 1B**). Six of the variants, p.(Arg39Cys), p.(Gln80Pro), p.(Ala315Ser), p.(Arg415Trp), p.(Arg429Trp), and p.(Arg498Trp) are located in the N-terminal coiled coil domain (**Figure 1B**). The variant p.(Ile870Asn) is located in the SAM1 domain (**Figure 1B**). The variants p.(Trp546Cys), p.(Arg784Trp), and p.(Ser906Leu) are in the disordered region of the protein, but p.(Ser906Leu) is located near the SAM1 domain (**Figure 1B**). Conservation analysis of the affected residues reveals that the residues in the coiled coil domain, p.Arg39, p.Gln80, p.Ala315, p.Arg415, and p.Arg429 are well-conserved in invertebrates and vertebrates (**Figure 1C**). The affected residue in the SAM1 domain, p.Ile870, and the affected residue near the SAM1 domain, p.Ser906, are also conserved across species. In the disordered region, p.Trp546 is conserved in the mice, but not in the fruit flies and worms. In contrast, p.Arg784 and p.Arg498Trp are conserved in mice and worms, but not in fruit flies. (**Figure 1C**).

Molecular modeling was completed for the missense variants using PyMol to determine if the amino acid changes affect the protein function *in silico* (**Figure 2**). Regarding the coiled coil variants, p.(Arg39Cys) removes the positively charged amino acid Arg and introduces an uncharged amino acid Cys (**Figure 2A**). Variant p.(Gln80Pro) introduces a ring structure (**Figure 2B**), and the variant p.(Ala315Ser) removes the hydrophobic amino acid and introduces a hydrophilic uncharged amino acid Ser (**Figure 2C**). The variant p.(Arg415Trp) introduces a bulky side chain predicted to disrupt the interaction with p.Gln411 (**Figure 2D**). The variant p.(Arg429Trp) introduces a bulky side chain that may disrupt the interaction with p.Asp424 (**Figure 2E**). The variant p.(Arg498Trp) introduces a bulky side chain (**Figure 2F**). In the disordered region, variant p.(Trp546Cys) results in the loss of the hydrophobic side chain (**Figure 2G**). The variant p.(Arg784Trp) introduces a bulky side chain predicted to disrupt the polar interaction with p.Asp785 (**Figure 2H**). Variant p.(Ile870Asn) is located in the SAM1 domain, and it replaces the hydrophobic amino acid Ile with the hydrophilic uncharged Asn (**Figure 2I**). The variant p.(Ser906Leu) is near the SAM1 domain and disrupts the interaction with the neighboring residue p.Ser908 (**Figure 2J**). Together, the molecular modeling suggests that these rare variants may hinder PPFIA3 protein function by disrupting the polar interactions.

**Figure 2:**
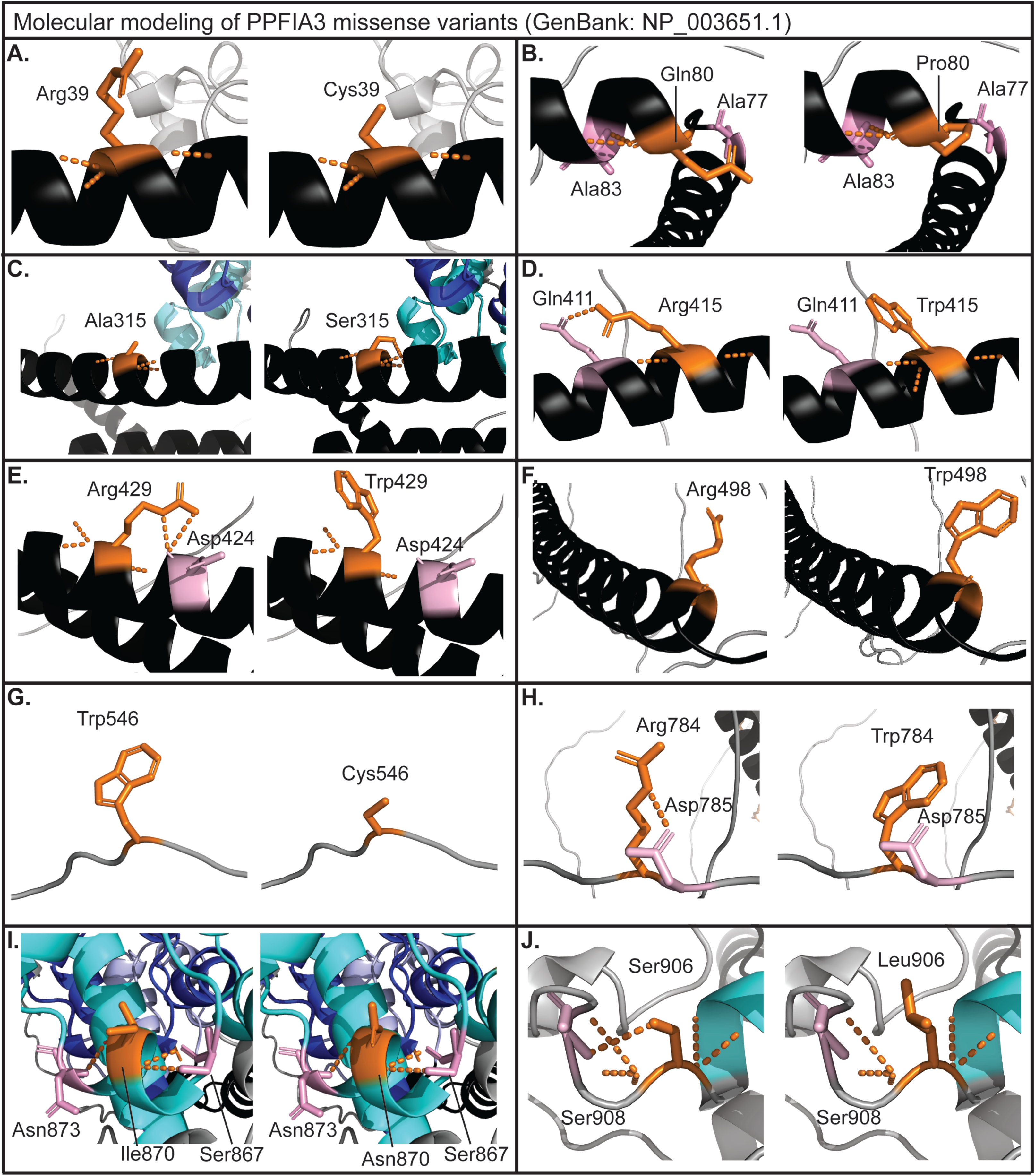
Molecular modeling of *PPFIA3* missense variants. (A-J) PPFIA3 missense variants are modeled in PyMol (version 2.5.2) with GenBank NP_003651.1. Human PPFIA3 WT residues are modeled in gray with coiled coils displayed in black and affected residues are highlighted in orange. Local polar contacts (orange dashed lines) and residue interactions (highlighted in pink) are displayed before and after mutagenesis for **(A)** p.(Arg39Cys), **(B)** p.(Gln80Pro), **(C)** p.(Ala315Ser), **(D)** p.(Arg415Trp), **(E)** p.(Arg429Trp), **(F)** p.(Arg498Trp), **(G)** p.(Trp546Cys), **(H)** p.(Arg784Trp), **(I)** p.(Ile870Asn), **(J)** p.(Ser906Leu).

### *In vivo* functional analysis of *PPFIA3* missense variants in fruit flies

To study the functional consequences of *PPFIA3* variants *in vivo*, we selected five of the missense variants to generate transgenic fruit flies using human cDNAs. We generated *UAS-PPFIA3-WT-HA, UAS-PPFIA3-p.(Arg39Cys)-HA, UAS-PPFIA3-p.(Ala315Ser)-HA, UAS-PPFIA3-p.(Arg415Trp)-HA, UAS-PPFIA3-p.(Trp546Cys)-HA,* and *UAS-PPFIA3-p.(Arg784Trp)-HA* fly alleles with C-terminal HA epitope tags. The GAL4-UAS expression system was used to express *PPFIA3* WT and variant cDNAs under the spatiotemporal regulation of the transactivator protein GAL4 (**Figure S1A**). A pan-neuronal driver on the second chromosome, elav-GAL4, was used to express *PPFIA3* cDNAs in neurons and a ubiquitous driver on the second chromosome, Actin-GAL4, was used to express *PPFIA3* cDNAs in the whole fly (**Figure S1A**). We verified elav-GAL4 mediated expression of PPFIA3 WT and variant proteins in 3^rd^ instar larval brains (**Figure S1B**). To confirm the cDNA copy number insertions, we isolated genomic DNA from adult flies from the following fly lines: *UAS-PPFIA3-WT-HA, UAS-PPFIA3-p.(Arg39Cys)-HA, UAS-PPFIA3-p.(Ala315Ser)-HA, UAS-PPFIA3-p.(Arg415Trp)-HA, UAS-PPFIA3-p.(Trp546Cys)-HA,* and *UAS-PPFIA3-p.(Arg784Trp)-HA*. Genomic DNA regions for *HA* epitope tag, *PPFIA3*, and *rps17* were amplified, and the PCR product band intensity of either *HA* or *PPFIA3* was quantified using *rps17* as the internal control (**Figures S1C, S2A-B**). No significance difference in the band intensity was observed, indicating the cDNA copy number is similar across all the *PPFIA3* WT and variant fly lines.

To determine if expression of *PPFIA3* WT and missense variants are deleterious to developmental processes, we ubiquitously expressed *PPFIA3* cDNAs using Actin-GAL4 and analyzed the fly developmental stages. *PPFIA3* cDNAs were expressed in the presence of endogenous fly *Liprin-*α. We found that expression of *PPFIA3* p.(Arg39Cys), p.(Ala315Ser), and p.(Arg415Trp) variants in the N-terminal coiled coil domain caused pupal lethality and eclosion defects, whereas this was not observed for the *PPFIA3* p.(Trp546Cys) and p.(Arg784Trp) variants in the disordered region (**Figure 3Ai-ii**). Although the expression of the coiled coil domain variants resulted in pupal lethality and eclosion defects, we obtained rare flies that eclosed (escapers). In these escapers, we observed a variable penetrance of leg dysmorphology. The wild-type morphology is comprised of three pairs of legs with each leg containing three segments: femur, tibia, and tarsus (**Figure 3Bi**). We found morphological defects in these segments in either the first, second, third, or all leg pairs with expression of the *PPFIA3* missense variants (**Figure 3Bi**). Leg dysmorphology was observed in 80% of *PPFIA3* p.(Arg39Cys) flies, 50% of *PPFIA3* p.(Ala315Ser) flies, 40% of *PPFIA3* p.(Arg415Trp) flies, and 10% of *PPFIA3* p.(Arg784Trp) flies (**Figure 3Bii**). In contrast, the leg dysmorphology phenotype was absent in the *PPFIA3* p.(Trp546Cys) flies (**Figure 3Bi-ii**).

**Figure 3:**
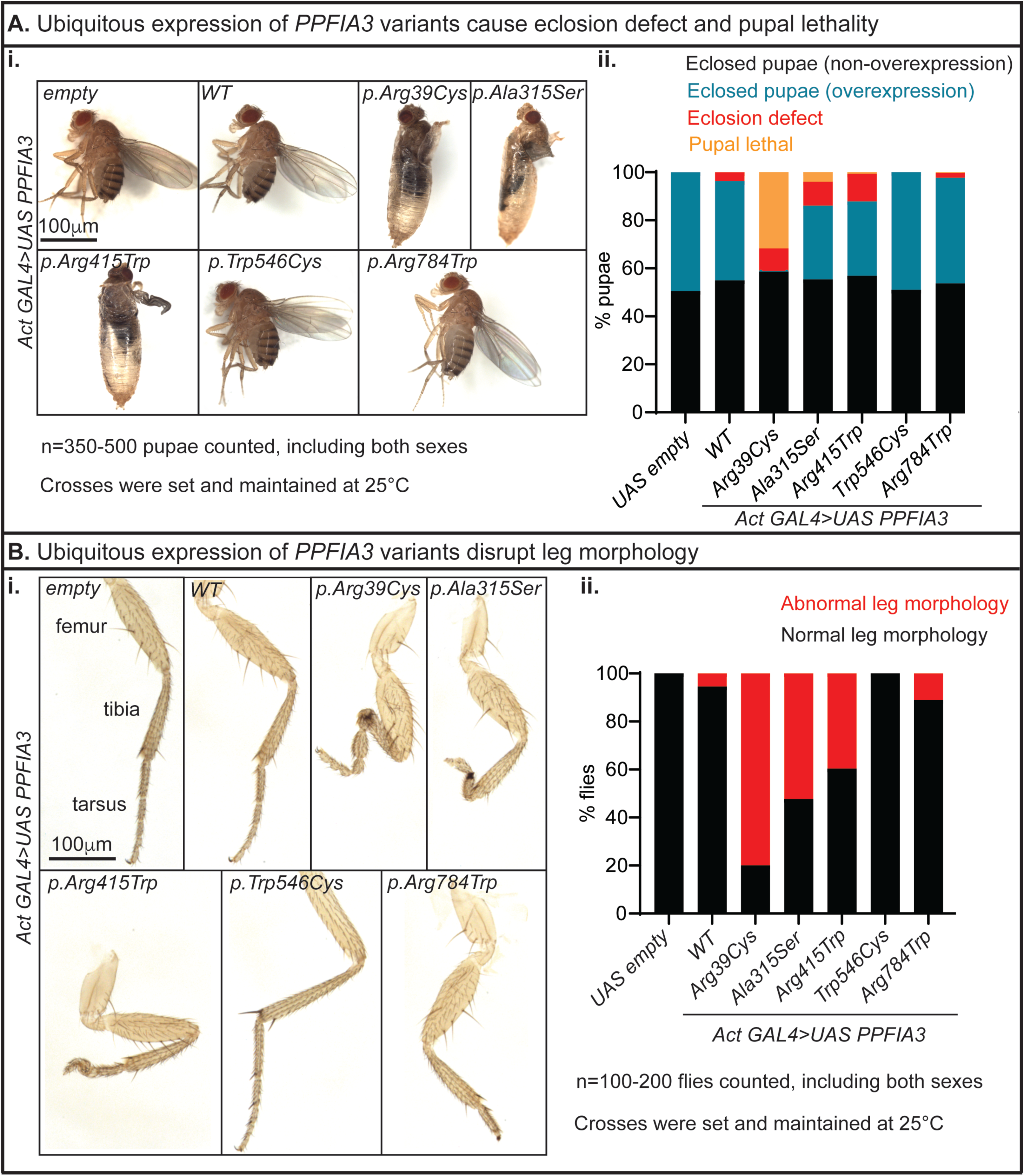
Act-GAL4 mediated ubiquitous expression of *PPFIA3* variants cause developmental and anatomical defects in fruit flies. (A) Pupal lethality and eclosion defect associated with Actin-GAL4-mediated overexpression of *PPFIA3* variants. Crosses were set and maintained at 25°C. **(i)** Representative images showing overexpression of *PPFIA3* p.(Arg39Cys), p.(Ala315Ser), and p.(Arg415Trp) cause pupal lethality and eclosion defect compared to the *PPFIA3* WT and *UAS-empty* control flies. Uneclosed flies from p.(Arg39Cys), p.(Ala315Ser), and p.(Arg415Trp) remain in the pupal case. Overexpression of *PPFIA3* p.(Trp546Cys) and p.(Arg784Trp) does not cause a difference in pupal lethality and eclosion defect compared to the *PPFIA3* WT and *UAS-empty* control flies. Scale bar is 100 μm. **(ii)** Quantification of pupal stage survival and eclosion defect from 350-500 pupae scored of both sexes. Black bars are % of pupae that eclose and do not express human PPFIA3 protein, blue bars are % of pupae that eclose and are overexpressing human PPFIA3 protein, red bars are % of pupae with eclosion defect, and orange bars are % of pupae with lethality. **(B)** Representative images of atypical leg morphology associated with Actin-GAL4-mediated overexpression of *PPFIA3* variants. Crosses were set and maintained at 25°C. **(i)** Empty control and PPFIA3 WT expressing flies have properly developed legs with three segments: femur, tibia, and tarsus. Expression of *PPFIA3* p.(Arg39Cys), p.(Ala315Ser), and p.(Arg415Trp) result in pronounced leg segment developmental defects compared to *PPFIA3* WT. Mild leg segmental developmental defects found with expression of *PPFIA3* p.(Arg784Trp) compared to *PPFIA3* WT. No leg segmental defects were found with expression of *PPFIA3* p.(Trp546Cys). Scale bar is 100 μm. **(ii)** Quantification of 100-200 flies of both sexes with red bars showing percent of flies with abnormal leg morphology and black bars showing percent of flies with normal leg morphology.

Next, to determine if the neuronal expression of *PPFIA3* variants by elav-GAL4 impaired nervous system development and function we conducted climbing behavior, bang sensitivity behavior, and NMJ morphology assays. First, we performed a climbing assay to assess for motor defects in negative geotaxis. The standard behavior of the flies is to climb upward, and any increase in time to climb represents a motor coordination defect. We found that *elav-GAL4>UAS-PPFIA3 WT* flies had motor function similar to control flies that do not express human PPFIA3 protein (*elav-GAL4 > UAS-empty*) (**Figure 4A**). However, climbing was impaired in both *elav-GAL4>UAS-PPFIA3 p.(Arg39Cys)* and *elav-GAL4>UAS-PPFIA3 p.(Arg415Trp)* expressing flies (**Figure 4A**). Second, to determine if the *PPFIA3* variants increase seizure susceptibility, we tested bang sensitivity in the flies. We found that *elav-GAL4>UAS-PPFIA3 WT* expressing flies were not bang sensitive and recovered similarly to the *elav-GAL4>UAS-empty* control. However, *elav-GAL4>UAS-PPFIA3 p.(Arg39Cys), elav-GAL4>UAS-PPFIA3 p.(Ala315Ser),* and *elav-GAL4>UAS-PPFIA3 p.(Arg415Trp)* flies exhibited bang sensitivity with an increased recovery time (**Figure 4B**). Third, to explore the consequence of *PPFIA3* variants at the synapse, we examined the fruit fly third instar larval NMJ morphology in muscle 6/7 of abdominal segment 3 (A3) (**Figure 4Ci-ii**). The fly NMJ is a glutamatergic synapse and a well-established model for excitatory glutamatergic synapse development and function.^40, 41^ We found reduced number of boutons (presynaptic contacts) with elav-GAL4 mediated expression of the *PPFIA3* p.(Arg39Cys) and p.(Arg415Trp) variants (**Figure 4Ciii**), indicating these variants perturb synapse formation. Total NMJ length associated with the *PPFIA3* variants remained similar to the *PPFIA3* WT and *UAS-empty* controls (**Figure 4Civ**).

**Figure 4:**
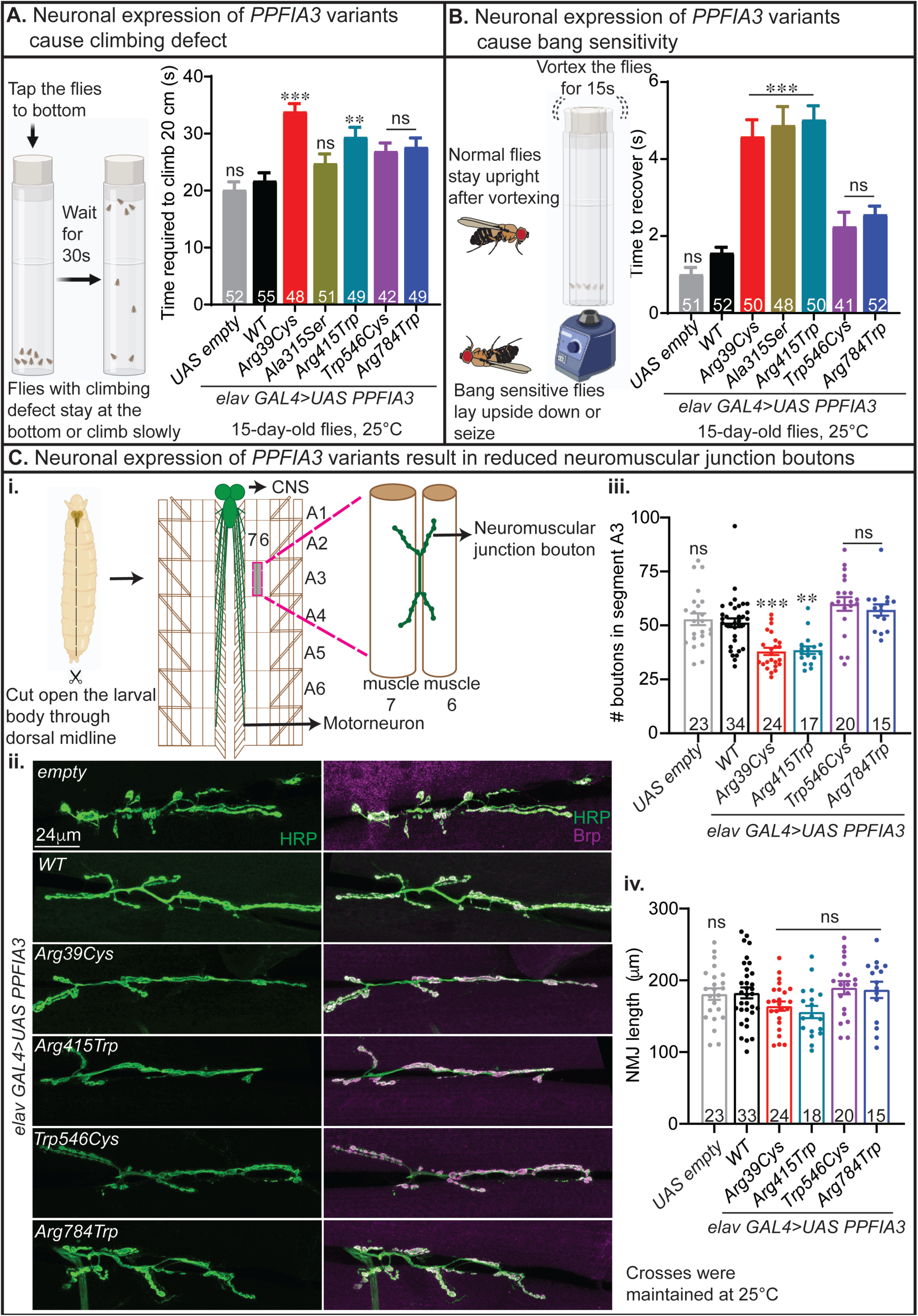
elav-GAL4 mediated neuronal overexpression of *PPFIA3* variants result in climbing defect, bang sensitivity, and neuromuscular junction (NMJ) bouton loss. **(A)** elav-GAL4 mediated neuronal expression of *PPFIA3* p.(Arg39Cys) and *PPFIA3* p.(Arg415Trp) result in impaired motor coordination on the climbing assay compared to *PPFIA3* WT and *empty* control flies. Crosses were set and maintained at 25°C. Behavioral testing was conducted at 20-21°C with both sexes. **(B)** elav-GAL4 mediated neuronal expression of *PPFIA3* p.(Arg39Cys), *PPFIA3* p.(Ala315Ser), and *PPFIA3* p.(Arg415Trp) have bang sensitivity with delayed recovery from vortexing compared to *PPFIA3* WT and *UAS-empty* control flies. Crosses were set and maintained at 25°C. Behavioral test was conducted at 20-21°C with both sexes. **(C)** elav-GAL4 mediated neuronal overexpression of *PPFIA3* variants result in NMJ bouton loss without a significant change in NMJ length. **(i)** Model depicting the method for visualizing the NMJ in fruit fly 3^rd^ instar larva. **(ii)** Representative images of 3^rd^ instar larval NMJs of each genotype including *elav GAL4>UAS empty, elav GAL4>PPFIA3 WT, p.(Arg39Cys), p.(Arg415Trp), p.(Trp546Cys),* and *p.(Arg784Trp)* are shown. HRP (Horseradish Peroxidase) is a pan-neuronal marker (green) and Brp (Bruchpilot) is an active zone marker (magenta). Scale bar is 24 μm. **(iii)** Quantification of total number of boutons in the muscle 6/7 (abdominal segment A3) NMJ show that the expression of *PPFIA3* p.(Arg39Cys) and p.(Arg415Trp) result in bouton loss compared to *PPFIA3* WT and empty control. In contrast, *PPFIA3* p.(Trp546Cys) and p.(Arg784Trp) show no alteration in bouton numbers. **(iv)** Quantification of total NMJ length in each genotype is shown and there is no significant difference between *PPFIA3* WT, variants, and *UAS-empty* control. Crosses were set and maintained at 25°C. Statistical analysis conducted with one-way ANOVA and Tukey’s post-hoc analysis. Data shown as mean ± SEM with the sample size of total number of quantified NMJs shown in the bars. Significance shown as \*\**p*<0.01, \*\*\**p*<0.001. Non-significance shown as ns.

To determine the functional nature of the human *PPFIA3* variants in the absence of wild-type fly *Liprin-*α, we performed *in vivo* rescue experiments with a previously established *Liprin-a* LOF allele^28^, *Liprin-*α*^F3ex15^*, and a *Liprin-a* deficiency allele, *Df(2L)Exel7027/CyO* (**Figure 5Ai**). First, we observed that complete loss of *Liprin*-α function (*Liprin-*α*^F3ex15^*/*Df(2L)Exel7027*) is embryonic lethal in control *da-GAL4>UAS-empty* expressing flies, with a few escapers reaching larval stage (**Figure 5Aii**). We expressed the human *PPFIA3* WT or variant cDNAs in the background of *Liprin*-α LOF using a ubiquitously expressed da-GAL4 at 20°C and assessed if expression of human PPFIA3 WT or variant proteins are able to rescue the embryonic lethality. We found a ∼25% larval rescue of embryonic lethality with *PPFIA3* WT, indicating functional conservation in fruit flies (**Figure 5Aii**). Expression of *PPFIA3* p.(Arg39Cys) and p.(Arg415Trp) resulted in significantly reduced larval rescue compared to WT (8% and 13%, respectively) (**Figure 5Aii**). However, expression of the *PPFIA3* p.(Trp546Cys) and p.(Arg784Trp) variants resulted in ∼17% rescue efficiency of the embryonic lethality, which was insignificant compared to *PPFIA3* WT. Second, we assessed the survival of the rescued larvae to the adult stage (**Figure 5Bi**). We found that ∼35% of *PPFIA3* WT expressing larvae reached the adult stage (**Figure 5Bii**), but none of the *PPFIA3* p.(Arg39Cys) expressing larvae reached the adult stage. In contrast, we found that 23% of *PPFIA3* p.(Arg415Trp) expressing larvae reached the adult stage, which is significantly reduced compared to *PPFIA3* WT (**Figure 5Bii**). However, the frequency of *PPFIA3* p.(Trp546Cys) and *PPFIA3* p.(Arg784Trp) expressing larvae reaching the adult stage was insignificant compared to *PPFIA3* WT (33% and 30%, respectively) (**Figure 5Bii**). Third, we assessed the survival of these rescue adults in the 48 hours post-eclosion. We found that <15% of *PPFIA3* WT rescue flies died, indicating expression of *PPFIA3* WT in the *Liprin-a* LOF background is capable of restoring viability and survival. In contrast, ∼60% of the *PPFIA3* p.(Arg415Trp) and 67% of the *PPFIA3* p.(Arg784Trp) rescue flies died within 48 hours post-eclosion (**Figure 5Biii**). However, *PPFIA3* p.(Trp546Cys) rescue flies had a survival rate similar to *PPFIA3* WT rescue flies (**Figure 5Biii**).

**Figure 5:**
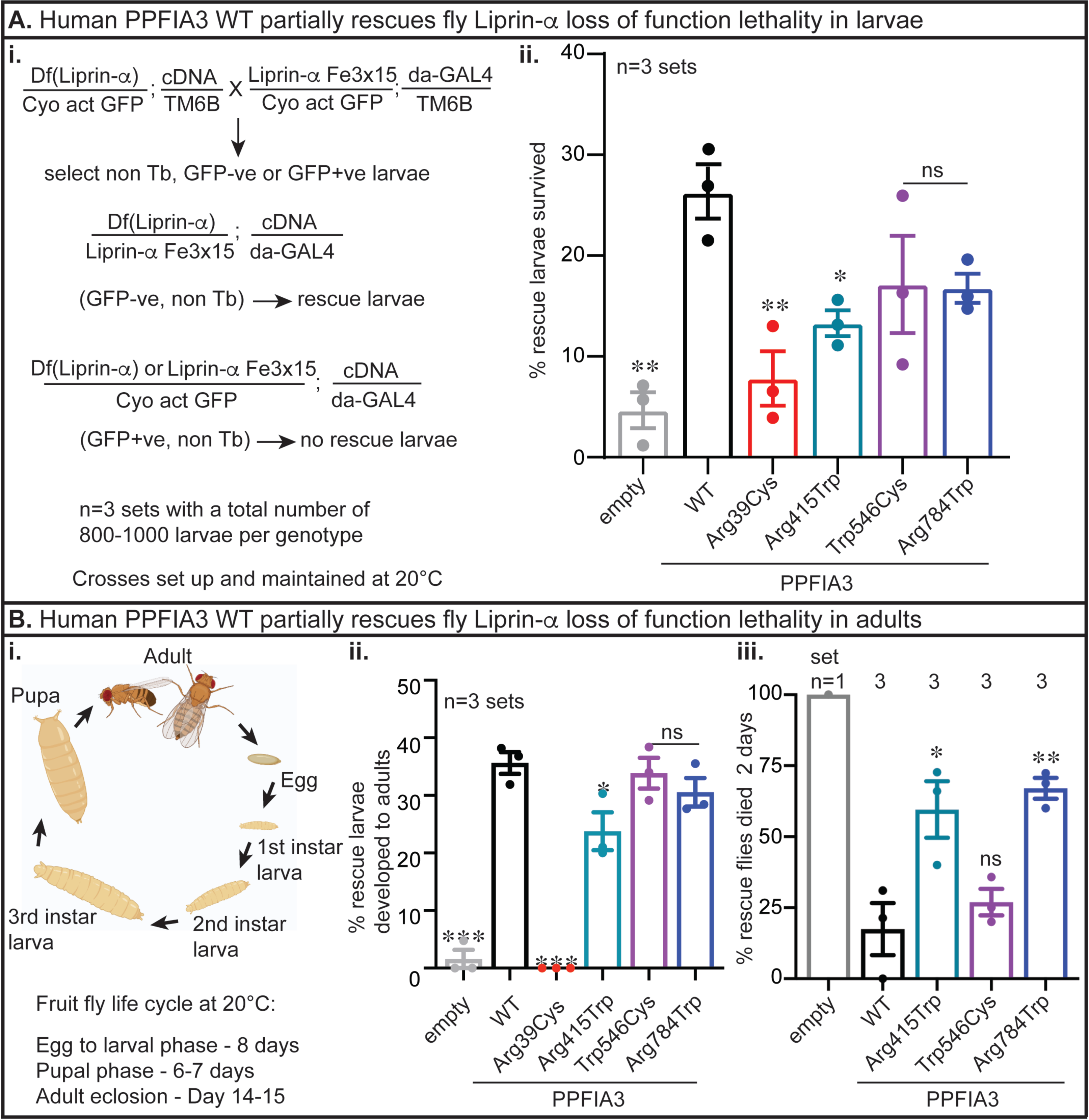
**PPFIA3 WT partially rescues the fly Liprin-**α **LOF lethality. (A)** Human PPFIA3 WT expression in the background of fly Liprin-a LOF results in a partial rescue of embryonic lethality. **(i)** Crossing-scheme to delete fly *Liprin-a* and express human *PPFIA3* WT and variants. The scheme describes the rescue larvae selection strategy. Crosses were set and maintained at 20°C. **(ii)** Quantification of n=800-1000 larvae per genotype showing % GFP-negative larvae (rescue larvae) that survive to the larval stage. *PPFIA3* WT expression is able to partially rescue larval viability compared to empty control. *PPFIA3* p.(Arg39Cys) and p.(Arg415Trp) show impaired ability to rescue larval viability. **(B)** Human *PPFIA3* WT expression partially rescues the lethality in adult stage. **(i)** Representative illustration of the different stages of fruit fly development. **(ii)** Quantification of 3 sets of rescued larvae per genotype that survive to the adult stage. *PPFIA3* WT expression is able to partially rescue adult viability compared to *empty* control. *PPFIA3* p.(Arg39Cys) and p.(Arg415Trp) show impaired ability to rescue adult viability. **(iii)** Quantification of 1-3 sets of rescued larvae per genotype that survive to 48 hours post-eclosion. For the empty control larvae only one escaper rescue larvae survived to adult stage, but died within 2 days post-eclosion. None of the *PPFIA3* p.(Arg39Cys) rescue larvae survived to adult stage. Due to the lack of any *PPFIA3* p.(Arg39Cys) rescue larvae surviving to the adult stage, this variant was not quantifiable for the adult survival phenotype. *PPFIA3* p.(Arg415Trp) and *PPFIA3* p.(Arg784Trp) show impaired ability to rescue adult viability compared to the *PPFIA3* WT. Statistical analysis with one-way ANOVA and Tukey’s post-hoc analysis. Data shown as mean ± SEM with the sample size of flies of both sexes scored shown in **(Ai)**. Significance shown as \**p*<0.05, \*\**p*<0.01, \*\*\**p*<0.001. Non-significance shown as ns.

Finally, we analyzed the number of NMJ boutons and NMJ length with da-GAL4 mediated expression of *PPFIA3* WT and variants in the *Liprin-*α LOF background at 20°C (**Figure S3A**). Although the total number of boutons are significantly reduced compared to *da-GAL4>UAS empty* controls, there is no significant difference in the number of boutons between *PPFIA3* WT and variants (**Figure S3Bi**). We quantified the total length of the NMJ and found no significant difference between genotypes (**Figure S3Bii**). Interestingly, we observed a significantly reduced ratio of bouton numbers per muscle 6/7 NMJ (segment A3) length in both *PPFIA3* WT and variants compared to the *da-GAL4>UAS empty* control (**Figure S3Bii**). However, the bouton to NMJ length ratio remained unchanged between *PPFIA3* WT and variants (**Figure S3Bii**). This indicates that there is a significant loss of bouton density in the background of complete *Liprin-a* LOF compared to the *da-GAL4>UAS empty* control. However, expression of neither *PPFIA3* WT nor variants were able to rescue the loss of NMJ boutons in the *Liprin-a* LOF background (**Figure S3Bi, iii**). It is possible that due to the severity of the complete *Liprin-a* LOF, the *PPFIA3* WT or variants expressing larvae examined for NMJ morphology represent a healthier subset of larvae capable of developing to the 3^rd^ instar stage. Therefore, we may not be capturing *PPFIA3* WT or variants expressing larvae with more severe NMJ phenotypes. This would limit our ability to identify a morphological difference between *PPFIA3* WT and variants in the background of complete *Liprin-a* LOF. Together, the *in vivo* fly functional experiments demonstrate that rare *PPFIA3* variants p.(Arg39Cys), p.(Arg415Trp), and p.(Arg784Trp) result in loss of PPFIA3 function and are deleterious to multiple developmental processes. These findings and our clinical characterizations show that rare *PPFIA3* mono-allelic variants in key functional domains lead to a syndromic neurodevelopmental disorder.

## Discussion

We describe 14 individuals with rare variants in *PPFIA3*, who have neurodevelopmental phenotypes including DD/ID, hypotonia, ASD, and epilepsy. The results of our clinical analysis, *in silico* molecular modeling, and *in vivo* functional studies in fruit flies show that rare *PPFIA3* variants lead to a previously unrecognized syndromic neurodevelopmental disorder. PPFIA3 protein domain analysis and molecular modeling revealed that six of the *PPFIA3* missense variants, p.(Arg39Cys), p.(Gln80Pro), p.(Ala315Ser), p.(Arg415Trp), p.(Arg429Trp), and p.(Arg498Trp) are located in the N-terminal coiled coil domain. The coiled coil domain is critical for PPFIA3’s homodimerization and interaction with active zone proteins, such as RIM and ELKS, to regulate active zone organization and synaptic vesicle release.^13–16^ Three *PPFIA3* missense variants, p.(Trp546Cys), p.(Arg784Trp), and p.(Ser906Leu), are located in the disordered region of the protein. One *PPFIA3* missense variant, p.(Ile870Asn), is located in the SAM1 domain. The SAM domains are known to bind to RNA and lipid membranes and the SAM domains of PPFIA proteins is shown to bind to the adhesion molecule LAR-RPTP.^17, 42, 43^ The *PPFIA3* frameshift deletion variant, p.(Glu1103Asnfs*8), may result in nonsense mediated decay followed by reduced protein expression.

Although interpretation is limited by the small sample size, we assessed the occurrence of six commonly reported features in the 14 individuals with rare *PPFIA3* mono-allelic variants and neurodevelopmental phenotypes. These six features include epilepsy or abnormal EEG, autism, DD/ID, hypotonia, dysmorphisms, and micro/macrocephaly (**Table 2**). We found that out of the six individuals with *PPFIA3* missense variants in the coiled coil domain (I:1, p.(Arg39Cys); I:2, p.(Gln80Pro); I:5, p.(Ala315Ser); I:6, p.(Arg415Trp); I:7, p.(Arg429Trp), and I:8 p.(Arg498Trp)), two of them had premature mortality (I:5 and I:7). I:1 and I:2 had 4/6 clinical features reported in which I:1 had epilepsy, DD/ID, hypotonia, and dysmorphisms and I:2 had autism, DD/ID, hypotonia, and dysmorphisms (**Table 2**). I:6 had 6/6 key clinical features reported with abnormal EEG, autism, DD/ID, hypotonia, dysmorphisms, and micro/macrocephaly (**Table 2**). Individual I:8 had 3/6 clinical features reported (autism, DD/ID, and dysmorphisms) (**Table 2**). Our amino acid conservation analysis showed that all affected residues in the coiled coil domain of PPFIA3 are highly conserved across mice, fruit flies, and worm, suggesting that these residues are critical for the protein’s function across different species. Molecular modeling of these variants suggested that changes in the affected residues could hinder the PPFIA3 protein function.

**Table 2:** Key clinical features in individuals with rare mono-allelic *PPFIA3* variants. The 14 *PPFIA3* variants identified in individuals I:1 - I:14 and corresponding locations in the protein domains are shown with the associated clinical features.

|  | PPFIA3 variants | Location | Key clinical features |  |  |  |  |  |  |  |
| --- | --- | --- | --- | --- | --- | --- | --- | --- | --- | --- |
|  |  |  | Epilepsy and/or abnormal EEG | Autism | DD/ID | Hypotonia | Dysmorphisms | Micro- or macrocephaly | Score (# present / total reported features) | Status |
| I:1 | c.115 C>T, p.(Arg39Cys) | Coiled coil | + | - | + | + | + | - | 4/6 | Alive |
| I:2 | c.239 A>C, p.(Gln80Pro) | Coiled coil | - | + | + | + | + | - | 4/6 | Alive |
| I:3 | c.240+1G>A | Intron | - | - | + | n/a | + | + | 3/5 | Alive |
| I:4 | c.240+1G>A | Intron | - | n/a | + | - | - | + | 2/5 | Alive |
| I:5 | c.943 G>T, p.(Ala315Ser) | Coiled coil | n/a | n/a | n/a | n/a | + | + | 2/2 | Deceased |
| I:6 | c.1243 C>T, p.(Arg415Trp) | Coiled coil | + | + | + | + | + | + | 6/6 | Alive |
| I:7 | c.1285 C>T, p.(Arg429Trp) | Coiled coil | n/a | n/a | n/a | n/a | n/a | n/a | n/a | Pregnancy terminated |
| I:8 | c.1492 C>T, p.(Arg498Trp) | Coiled coil | - | + | + | - | + | - | 3/6 | Alive |
| I:9 | c.1638 G>T, p.(Trp546Cys) | Disordered region | - | + | + | - | - | - | 2/6 | Alive |
| I:10 | c.2350 C>T, p.(Arg784Trp) | Disordered region | + | - | + | - | - | - | 2/6 | Alive |
| I:11 | c.2609 T>A, p.(Ile870Asn) | SAM1 | - | + | + | - | - | + | 3/6 | Alive |
| I:12 | c.2609 T>A, p.(Ile870Asn) | SAM1 | - | - | + | + | - | + | 3/6 | Alive |
| I:13 | c.2717 C>T, p.(Ser906Leu) | Disordered region | + | + | + | + | + | + | 6/6 | Alive |
| I:14 | c.3307del, p.(Glu1103Asnfs*8) | SAM3 | + | - | + | + | + | - | 4/6 | Alive |
| Abbreviations: electroencephalogram (EEG), delayed development (DD), intellectual disability (ID), sterile alpha motif (SAM), no information available (n/a) |  |  |  |  |  |  |  |  |  |  |

Three individuals have *de novo PPFIA3* missense variants (I:9, p.(Trp546Cys); I:10, p.(Arg784Trp); and I:13, p.(Ser906Leu)) in the disordered region of the protein. The amino acid conservation analysis suggests that residues in the disordered region of PPFIA3 are not well conserved across species except for p.Ser906 that is near the SAM1 domain. Individuals I:9 (p.(Trp546Cys)) and I:10 (p.(Arg784Trp)) has 2/6 clinical features reported (**Table 2**). I:9 had autism and DD/ID, whereas I:10 had epilepsy and DD/ID. The small number of phenotypic findings in I:9 and I:10 may be due to the location of the variants in a disordered region of the PPFIA3 protein. Individual I:13 (p.(Ser906Leu)) had 6/6 clinical features reported with abnormal EEG, autism, DD/ID, hypotonia, dysmorphisms, and micro/macrocephaly. Together, these findings suggest that variants in the disordered regions are associated with variable disease severity, which may be due to the lack of association in this region with functional domains for PPFIA3. Two individuals had *PPFIA3* missense variants in the SAM1 or SAM3 domains (I:11, p.(Ile870Asn); I:12 p.(Ile870Asn)). Individuals I:11 and I:12 are monozygotic twins and both had 3/6 clinical features reported. I:11 had autism, DD/ID, and microcephaly. I:12 had DD/ID, hypotonia, and microcephaly. One individual has a *PPFIA3* frameshift variant in the SAM3 domain, I:14, p.(Glu1103Asnfs*8), with 4/6 clinical features reported that include abnormal EEG, DD/ID, hypotonia, and dysmorphisms. There were two individuals (I:3, I:4) with the same *PPFIA3* intronic splice variant (c.240+1G>A) and the number of key clinical features reported were 3/5 (I:3) and 2/5 (I:4). Individual I:3 inherited the *PPFIA3* variant from the affected parent, I:4. The inheritance of the *PPFIA3* variant in I:4 is unknown. Both I:3 and I:4 have DD, ID, and microcephaly. Finally, we also identified a missense *PPFIA3* p.(Arg559Trp) variant of unknown inheritance in I:15. The severe neurodegeneration phenotype was not observed in the 14 individuals in our cohort, suggesting that other genetic alterations or etiologies may contribute to I:15’s clinical findings.

Our conservation analysis reveal that the PPFIA3 protein domains are well conserved in the fruit fly homolog, Liprin-α, which is primarily expressed in the fruit fly embryonic and larval nervous system.^28^ In the fruit fly larvae, it is found in the NMJ synapses.^28^ Several previous fruit fly studies show that Liprin-α is critical for synapse morphogenesis and axon guidance with Liprin-α LOF resulting in a reduced number of boutons, axon branching, and active zone dimensions.^28^ Another important function of Liprin-α is the regulation of synaptic vesicle trafficking. Liprin-α LOF causes an accumulation of synaptic markers in the axon with a decrease in anterograde transport and an increase in retrograde transport.^44^ Moreover, Liprin-α is also essential for retinal axon targeting in fruit flies.^30^ Interestingly, a previous study with *Ppfia3* knockout mice revealed that complete loss of *Ppfia3* impairs synaptic vesicle exocytosis, reduced the number of synapses that were responsive to action potentials, and altered the active zone structure.^24^ Double knockout of mouse *Ppfia2* and *Ppfia3* in hippocampal neurons impaired neurotransmitter release due to perturbations in the number of docked vesicles per synapse.^45^ Together, these findings reveal that *PPFIA3* homologs in fruit flies and mice are crucial for multiple neurodevelopmental processes.

We found that the ubiquitous expression of *PPFIA3* missense variants in the coiled coil domain (p.(Arg39Cys), p.(Ala315Ser), and p.(Arg415Trp)) resulted in pupal lethality, eclosion defects, and abnormal leg morphology. Neuron-specific expression revealed seizure-like behaviors and climbing defects with *PPFIA3* p.(Arg39Cys) and p.(Arg415Trp) variants. As Liprin-α is known to regulate NMJ development in fruit flies, we examined the effect of human *PPFIA3* WT and variant cDNA expression in the larval NMJ. We found that overexpression of *PPFIA3* p.(Arg39Cys) and p.(Arg415Trp) caused a reduction in bouton number compared to *PPFIA3* WT, which would potentially impair neurotransmission. Our fly overexpression findings reveal the *PPFIA3* missense variants in the coiled coil domains cause lethality, suggesting these variants are dominant negative alleles. Interestingly, ubiquitous, or neuronal overexpression of the *PPFIA3* variants in the disordered region (p.(Arg784Trp) and p.(Trp546Cys)) had either mild or no phenotypes in the fruit flies, which may be related to the fact that the variants are either not conserved in the flies or cause mild protein dysfunction that is tolerated in the fly model.

To further determine whether the variants are LOF or gain-of-function (GOF) in nature and the functional conservation between human *PPFIA3* and fly *Liprin-*α, we performed a LOF lethality rescue assay using *Liprin-*α mutants and *PPFIA3* WT and variants. Our LOF rescue assay showed that human *PPFIA3* WT expression partially rescued the *Liprin-*α LOF embryonic lethality and development to the adult stage, suggesting that human PPFIA3 WT protein function is partially conserved in fruit flies. Interestingly, expression of the coiled coil variants, *PPFIA3* p.(Arg39Cys) and p.(Arg415Trp) showed reduced rescue efficiency of the *Liprin-*α LOF embryonic lethality, indicating these variants are strong LOF variants. In contrast, expression of the disordered region variants, *PPFIA3* p.(Trp546Cys) and p.(Arg784Trp) showed a similar rescue efficiency of the embryonic lethality as compared to *PPFIA*3 WT expression. However, we found in the adult stage that *PPFIA3* p.(Arg784Trp) expression significantly reduced the lifespan of rescue flies compared to *PPFIA3* WT, suggesting that p.(Arg784Trp) is a hypomorphic LOF variant. These findings are consistent with the clinical findings in individual I:9, p.(Trp546Cys) and I:10, p.(Arg784Trp). Both individuals had only 2/6 commonly reported features (**Table 2**). In contrast, individuals I:1, p.(Arg39Cys) and I:6, p.(Arg415Trp) had 4/6 and 6/6 commonly reported features, respectively (**Table 2**). Together, our overexpression and LOF rescue assays in fruit flies reveal that rare mono-allelic *PPFIA3* variants cause a neurodevelopmental disorder through a LOF mechanism and that disease severity correlates with the degree of LOF. The clinical phenotypes and functional assays in fruit flies (**Table 3**) point towards a possible domain-specific disease severity mechanism where the variants in the coiled coil domains might lead to relatively more severe phenotypes in both affected individuals and fruit flies.

**Table 3:** Comparison of clinical phenotypes and findings in fruit flies expressing *PPFIA3* variants. The five *PPFIA3* missense variants (I:1, p.(Arg39Cys); I:5, p.(Ala315Ser); I:6, p.(Arg415Trp); I:9, p.(Trp546Cys); I:10, p.(Arg784Trp)) and their corresponding locations in the protein domains are shown with the associated clinical features and summary of functional analysis findings in the fruit fly assays.

|  | Individual | I:1 | I:5 | I:6 | I:9 | I:10 |
| --- | --- | --- | --- | --- | --- | --- |
|  | *PPFIA3* variants | c.115 C>T<br>(p.Arg39Cys) | c.943 G>T<br>(p.Ala315Ser) | c.1243 C>T<br>(p.Arg415Trp) | c.1638 G>T<br>(p.Trp546Cys) | c.2350 C>T<br>(p.Arg784Trp) |
| Key clinical phenotypes in individuals with *PPFIA3* variants | Location | Coiled coil | Coiled coil | Coiled coil | Disordered region | Disordered region |
|  | Epilepsy and/or abnormal EEG | + | n/a | + | - | + |
|  | Autism | - | n/a | + | + | - |
|  | DD/ID | + | n/a | + | + | + |
|  | Hypotonia | + | n/a | + | - | - |
|  | Dysmorphisms | + | + | + | - | - |
|  | Micro- or macrocephaly | - | + | + | - | - |
|  | Score (# present / total reported features) | 4/6 | 2/2 | 6/6 | 2/6 | 2/6 |
| Findings in fruit flies expressing *PPFIA3* variants | Eclosion defect | + | + | + | - | - |
|  | Abnormal leg morphology | + | + | + | - | + |
|  | Climbing defect | + | - | + | - | - |
|  | Bang sensitivity | + | + | + | - | - |
|  | NMJ defect | + | n/a | + | - | - |
|  | Liprin-a LOF rescue | + | n/a | + | - | + |
|  | Score (# present / total assays) | 6/6 | 3/4 | 6/6 | 0/6 | 2/6 |

**Abbreviations:** electroencephalogram (EEG), delayed development (DD), intellectual disability (ID), no information available (n/a), loss of function (LOF), neuromuscular junction (NMJ)

In summary, our study provides clinical and functional evidence that rare mono-allelic *PPFIA3* variants cause a syndromic neurodevelopmental disorder characterized by DD/ID, hypotonia, autism, and epilepsy. Our *in vivo* functional modeling in fruit flies reveal that *PPFIA3* variants may contribute to disease pathogenesis through LOF mechanisms related to the location of the affected residues in the PPFIA3 functional domains. Analysis of potential genotype-phenotype correlations using the clinical and *in vivo* fruit fly findings suggest that *PPFIA3* variants in the coiled coil domains may have relatively more deleterious effects. A longitudinal assessment in a larger sample size of affected individuals and functional studies in model organisms would advance our understanding of disease pathogenesis, improve prognostication based on variant type and location, and identify potential therapeutic avenues.

## Data Availability

All data produced in the present study are available upon reasonable request to the authors. The accession numbers for the variants reported to ClinVar are (1) I:1, ClinVar: SUB12878798; GenBank:NM_003660.4 (PPFIA3); c.115 C>T (p.Arg39Cys); (2) I:2, ClinVar: SUB12879592; GenBank:NM_003660.4 (PPFIA3); c.239 A>C (p.Gln80Pro); (3) I:3, ClinVar: SUB12866061; GenBank:NM_003660.4 (PPFIA3); c.240+1G>A; (4) I:4, ClinVar: SUB12866110; GenBank:NM_003660.4 (PPFIA3); c.240+1G>A; (5) I:6, ClinVar: SUB12879585; GenBank:NM_003660.4 (PPFIA3); c.1243 C>T (p.Arg415Trp); (6) I:7, ClinVar: SUB12690823; GenBank: NM_003660.4 (PPFIA3); c.1285C>T (p.Arg429Trp); (7) I:8, ClinVar: SUB12947536; GenBank:NM_003660.4 (PPFIA3); c.1492 C>T (p.Arg498Trp); (8) I:9, ClinVar: SUB12879586; GenBank:NM_003660.4 (PPFIA3); c.1638 G>T (p.Trp546Cys); (9) I:10, ClinVar: SUB12689969; GenBank: NM_003660.4 (PPFIA3); c.2350 C>T (p.Arg784Trp); (10) I:11 and I:12, ClinVar: SUB12879588; GenBank:NM_003660.4 (PPFIA3); c.2609T>A (p.Ile870Asn); (11) I:13, ClinVar: SCV003035511; GenBank: NM_003660.4 (PPFIA3); c.2717 C>T (p.Ser906Leu); (12) I:14, ClinVar: SCV003035512; GenBank: NM_003660.4 (PPFIA3); c.3307del (p.Glu1103Asnfs*8); (13) I:15, ClinVar: SUB12693099; GenBank: NM_003660.4 (PPFIA3); c.1675C>T (p.Arg559Trp)

## Acknowledgements

We thank the families and clinical staff at each location for participation in this study. We thank the CCuB for technical support and management of the computing platform for the individuals I:3 and I:4. In addition, we thank Mingshan Xue, Dongwon Lee, Kailin Mao, Wu (Charles) Chen, Brooke Horist, and Cole Deisseroth for critical feedback on the manuscript.

## Data availability

The accession numbers for the variants reported to ClinVar are (1) I:1, ClinVar: SUB12878798; GenBank:NM_003660.4 (PPFIA3); c.115 C>T (p.Arg39Cys); (2) I:2, ClinVar: SUB12879592; GenBank:NM_003660.4 (PPFIA3); c.239 A>C (p.Gln80Pro); (3) I:3, ClinVar: SUB12866061; GenBank:NM_003660.4 (PPFIA3); c.240+1G>A; (4) I:4, ClinVar: SUB12866110; GenBank:NM_003660.4 (PPFIA3); c.240+1G>A; (5) I:6, ClinVar: SUB12879585; GenBank:NM_003660.4 (PPFIA3); c.1243 C>T (p.Arg415Trp); (6) I:7, ClinVar: SUB12690823; GenBank: NM_003660.4 (PPFIA3); c.1285C>T (p.Arg429Trp); (7) I:8, ClinVar: SUB12947536; GenBank:NM_003660.4 (PPFIA3); c.1492 C>T (p.Arg498Trp); (8) I:9, ClinVar: SUB12879586; GenBank:NM_003660.4 (PPFIA3); c.1638 G>T (p.Trp546Cys); (9) I:10, ClinVar: SUB12689969; GenBank: NM_003660.4 (PPFIA3); c.2350 C>T (p.Arg784Trp); (10) I:11 and I:12, ClinVar: SUB12879588; GenBank:NM_003660.4 (PPFIA3); c.2609T>A (p.Ile870Asn); (11) I:13, ClinVar: SCV003035511; GenBank: NM_003660.4 (PPFIA3); c.2717 C>T (p.Ser906Leu); (12) I:14, ClinVar: SCV003035512; GenBank: NM_003660.4 (PPFIA3); c.3307del (p.Glu1103Asnfs*8); (13) I:15, ClinVar: SUB12693099; GenBank: NM_003660.4 (PPFIA3); c.1675C>T (p.Arg559Trp)

## Supplementary material

Supplementary material includes seven tables and three figures.

## Appendix 1

**Members of the Undiagnosed Diseases Network (UDN):** Maria T. Acosta, Margaret Adam, David R. Adams, Raquel L. Alvarez, Justin Alvey, Laura Amendola, Ashley Andrews, Euan A. Ashley, Carlos A. Bacino, Guney Bademci, Ashok Balasubramanyam, Dustin Baldridge, Jim Bale, Michael Bamshad, Deborah Barbouth, Pinar Bayrak-Toydemir, Anita Beck, Alan H. Beggs, Edward Behrens, Gill Bejerano, Hugo J. Bellen, Jimmy Bennett, Beverly Berg-Rood, Jonathan A. Bernstein, Gerard T. Berry, Anna Bican, Stephanie Bivona, Elizabeth Blue, John Bohnsack, Devon Bonner, Lorenzo Botto, Brenna Boyd, Lauren C. Briere, Gabrielle Brown, Elizabeth A. Burke, Lindsay C. Burrage, Manish J. Butte, Peter Byers, William E. Byrd, John Carey, Olveen Carrasquillo, Thomas Cassini, Ta Chen Peter Chang, Sirisak Chanprasert, Hsiao-Tuan Chao, Ivan Chinn, Gary D. Clark, Terra R. Coakley, Laurel A. Cobban, Joy D. Cogan, Matthew Coggins, F. Sessions Cole, Heather A. Colley, Heidi Cope, Rosario Corona, William J. Craigen, Andrew B. Crouse, Michael Cunningham, Precilla D’Souza, Hongzheng Dai, Surendra Dasari, Joie Davis, Jyoti G. Dayal, Esteban C. Dell’Angelica, Katrina Dipple, Daniel Doherty, Naghmeh Dorrani, Argenia L. Doss, Emilie D. Douine, Dawn Earl, David J. Eckstein, Lisa T. Emrick, Christine M. Eng, Marni Falk, Elizabeth L. Fieg, Paul G. Fisher, Brent L. Fogel, Irman Forghani, William A. Gahl, Ian Glass, Bernadette Gochuico, Page C. Goddard, Rena A. Godfrey, Katie Golden-Grant, Alana Grajewski, Don Hadley, Sihoun Hahn, Meghan C. Halley, Rizwan Hamid, Kelly Hassey, Nichole Hayes, Frances High, Anne Hing, Fuki M. Hisama, Ingrid A. Holm, Jason Hom, Martha Horike-Pyne, Alden Huang, Sarah Hutchison, Wendy Introne, Rosario Isasi, Kosuke Izumi, Fariha Jamal, Gail P. Jarvik, Jeffrey Jarvik, Suman Jayadev, Orpa Jean-Marie, Vaidehi Jobanputra, Lefkothea Karaviti, Shamika Ketkar, Dana Kiley, Gonench Kilich, Shilpa N. Kobren, Isaac S. Kohane, Jennefer N. Kohler, Susan Korrick, Mary Kozuira, Deborah Krakow, Donna M. Krasnewich, Elijah Kravets, Seema R. Lalani, Byron Lam, Christina Lam, Brendan C. Lanpher, Ian R. Lanza, Kimberly LeBlanc, Brendan H. Lee, Roy Levitt, Richard A. Lewis, Pengfei Liu, Xue Zhong Liu, Nicola Longo, Sandra K. Loo, Joseph Loscalzo, Richard L. Maas, Ellen F. Macnamara, Calum A. MacRae, Valerie V. Maduro, AudreyStephannie Maghiro, Rachel Mahoney, May Christine V. Malicdan, Laura A. Mamounas, Teri A. Manolio, Rong Mao, Kenneth Maravilla, Ronit Marom, Gabor Marth, Beth A. Martin, Martin G. Martin, Julian A. Martínez-Agosto, Shruti Marwaha, Jacob McCauley, Allyn McConkie-Rosell, Alexa T. McCray, Elisabeth McGee, Heather Mefford, J. Lawrence Merritt, Matthew Might, Ghayda Mirzaa, Eva Morava, Paolo Moretti, John Mulvihill, Mariko Nakano-Okuno, Stanley F. Nelson, John H. Newman, Sarah K. Nicholas, Deborah Nickerson, Shirley Nieves-Rodriguez, Donna Novacic, Devin Oglesbee, James P. Orengo, Laura Pace, Stephen Pak, J. Carl Pallais, Christina G.S. Palmer, Jeanette C. Papp, Neil H. Parker, John A. Phillips III, Jennifer E. Posey, Lorraine Potocki, Barbara N. Pusey Swerdzewski, Aaron Quinlan, Deepak A. Rao, Anna Raper, Wendy Raskind, Genecee Renteria, Chloe M. Reuter, Lynette Rives, Amy K. Robertson, Lance H. Rodan, Jill A. Rosenfeld, Natalie Rosenwasser, Francis Rossignol, Maura Ruzhnikov, Ralph Sacco, Jacinda B. Sampson, Mario Saporta, Judy Schaechter, Timothy Schedl, Kelly Schoch, Daryl A. Scott, C. Ron Scott, Elaine Seto, Vandana Shashi, Jimann Shin, Edwin K. Silverman, Janet S. Sinsheimer, Kathy Sisco, Edward C. Smith, Kevin S. Smith, Lilianna Solnica-Krezel, Ben Solomon, Rebecca C. Spillmann, Joan M. Stoler, Kathleen Sullivan, Jennifer A. Sullivan, Angela Sun, Shirley Sutton, David A. Sweetser, Virginia Sybert, Holly K. Tabor, Queenie K.-G. Tan, Amelia L. M. Tan, Arjun Tarakad, Mustafa Tekin, Fred Telischi, Willa Thorson, Cynthia J. Tifft, Camilo Toro, Alyssa A. Tran, Rachel A. Ungar, Tiina K. Urv, Adeline Vanderver, Matt Velinder, Dave Viskochil, Tiphanie P. Vogel, Colleen E. Wahl, Melissa Walker, Stephanie Wallace, Nicole M. Walley, Jennifer Wambach, Jijun Wan, Lee-kai Wang, Michael F. Wangler, Patricia A. Ward, Daniel Wegner, Monika Weisz Hubshman, Mark Wener, Tara Wenger, Monte Westerfield, Matthew T. Wheeler, Jordan Whitlock, Lynne A. Wolfe, Kim Worley, Changrui Xiao, Shinya Yamamoto, John Yang, Zhe Zhang, Stephan Zuchner

---

**Figure S1: GAL4-UAS targeted expression of PPFIA3 WT and variant cDNAs in fruit flies. (A)** Methodology for neuron specific and ubiquitous expression of *UAS-PPFIA3*-WT and variant cDNAs using elav-GAL4 and Actin-GAL4, respectively. **(B)** elav-GAL4 mediated expression of *PPFIA3* WT and five missense variants are shown in the third instar larval brain using immunostaining against the C-terminal HA epitope tag (magenta). Nuclear staining with DAPI is shown in cyan. Scale bar is 60 μm. Crosses were set and maintained at 25°C. **(C) (i)** Representative PCR from genomic DNA showing the relative *HA* and *PPFIA3* levels in *UAS-PPFIA3* WT or variant fly lines relative to the endogenous gene *rps17*. No differences in **(ii)** *HA* and **(iii)** *PPFIA3* levels were observed between the *PPFIA3* variants compared to *PPFIA3* WT. Statistical analysis with one-way ANOVA and Tukey’s post-hoc test. Data shown as mean ± standard error of mean (SEM).

**Figure S2: Original genomic PCR DNA gel images. (A)** Raw image of genomic PCR results from Set 1 used for representative image and quantification in Figure S1. **(B)** Raw image of genomic PCR results from Sets 2 and 3 used for quantification in Figure S1.

**Figure S3: da-GAL4 mediated ubiquitous expression of PPFIA3 WT and variant cDNAs in the background of complete loss of *Liprin-a* result in NMJ bouton loss. (A)** Representative images of 3^rd^ instar larval NMJs of each genotype including *da-GAL4>UAS empty, da GAL4>PPFIA3 WT, p.(Arg39Cys), p.(Ala315Ser), p.(Trp546Cys),* and *p.(Arg784Trp)* in the complete loss of *Liprin-a* background is shown. HRP (Horseradish Peroxidase) is a pan-neuronal marker (green) and Brp (Bruchpilot) is an active zone marker (magenta). Scale bar is 24 μm. **(B) (i)** Quantification of total number of boutons in the muscle 6/7 (abdominal segment A3) NMJ show that the expression of *PPFIA3* WT and variants in the background of *Liprin-a* LOF results in significant loss of boutons compared to the *da-GAL4>UAS empty* control. However, there is no significant difference in the NMJ bouton numbers between *PPFIA3* WT and variants. **(ii)** Quantification of total NMJ length show no significant difference. **(iii)** Quantification of the ratio of boutons per NMJ length show a significance difference between *da-GAL4>UAS empty* control and other genotypes, but there is no significant difference between *PPFIA3* WT and variants. Statistical analysis with one-way ANOVA and Tukey’s post-hoc analysis. Data shown as mean ± SEM with the sample size of total number of NMJs shown in figure. Significance shown as \*\*\**p*<0.001. Non-significance shown as ns.

**Table S1: Primer sequences for Q5 mutagenesis, sequencing, and genomic PCR.**

**Table S2: *In silico* predictions for *PPFIA3* variants (individuals I:1 to I:14).**

**Table S3: Clinical and genetic findings in individual I:15 with discordant neurodegenerative phenotype.**

**Table S4: General information and clinical findings in individuals I:1 - I:7 with *PPFIA3* variants.**

**Table S5: General information and clinical findings in individuals I:8 - I:14 with *PPFIA3* variants.**

**Table S6: MRI brain findings in individuals I:1 - I:14 with *PPFIA3* variants.**

**Table S7: Statistical summary of data for *PPFIA3* variant studies.**

