## Supplementary material for "Rare variants in *PPFIA3* cause delayed development, intellectual disability, autism, and epilepsy"

Figure S1

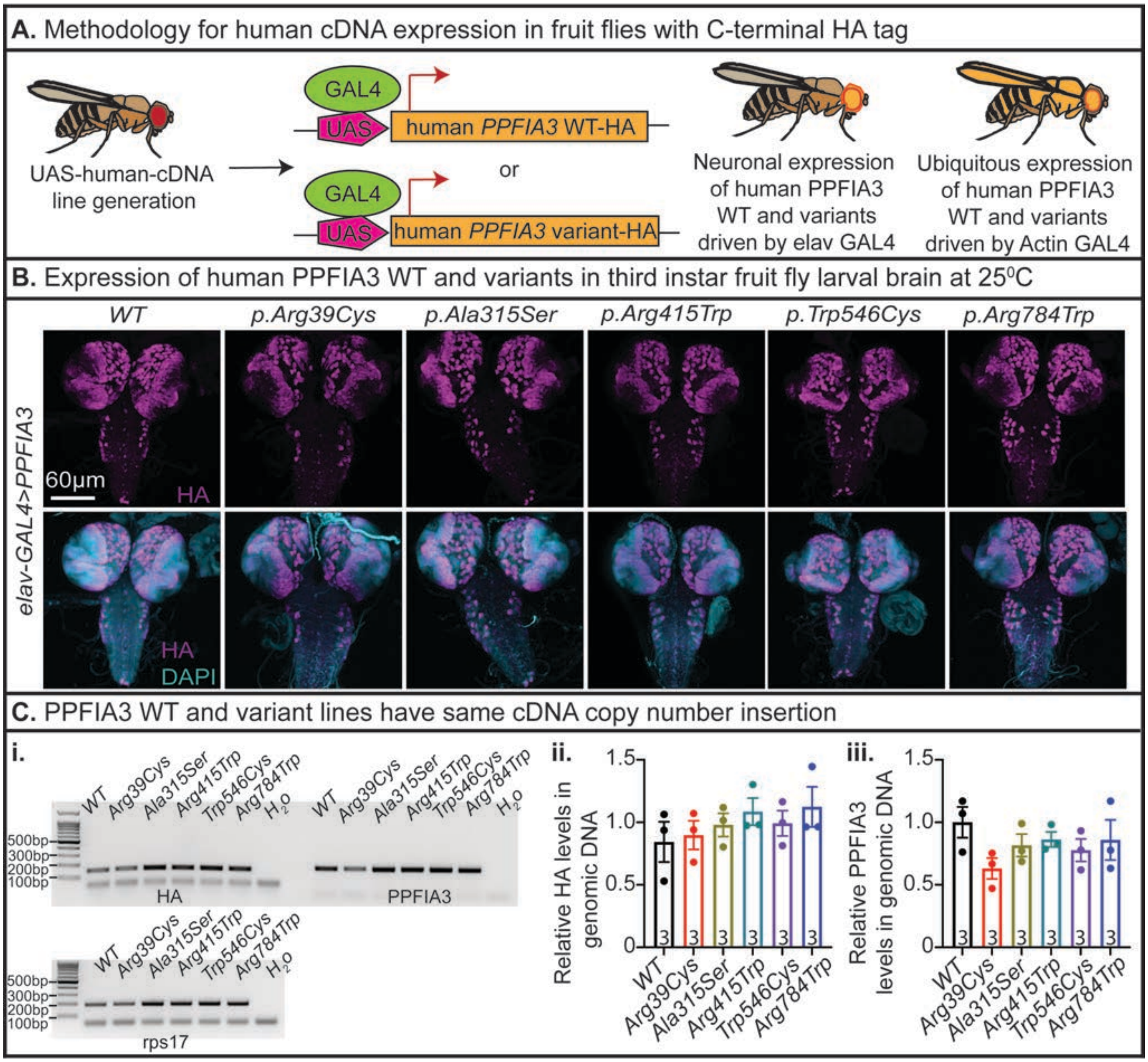

A. Methodology for human cDNA expression in fruit flies with C-terminal HA tag
GAL4
UAS
human PPFIA3 WT-HA
or
human PPFIA3 variant-HA
UAS-human-cDNA line generation
Neuronal expression of human PPFIA3 WT and variants driven by elav GAL4
Ubiquitous expression of human PPFIA3 WT and variants driven by Actin GAL4
B. Expression of human PPFIA3 WT and variants in third instar fruit fly larval brain at 25⁰C
WT
p.Arg39Cys
p.Ala315Ser
p.Arg415Trp
p.Trp546Cys
p.Arg784Trp
elav-GAL4>PPFIA3
60µm
HA
HA
DAPI
C. PPFIA3 WT and variant lines have same cDNA copy number insertion
i.
WT
Arg39Cys
Ala315Ser
Arg415Trp
Trp546Cys
Arg784Trp
H2O
500bp
300bp
200bp
100bp
HA
PPFIA3
rps17
ii.
Relative HA levels in genomic DNA
iii.
Relative PPFIA3 levels in genomic DNA
1.5
1.0
0.5
0
3

Figure S2

**A.** Set 1 genomic DNA PCR raw image for representative image and quantifications in Figure S2

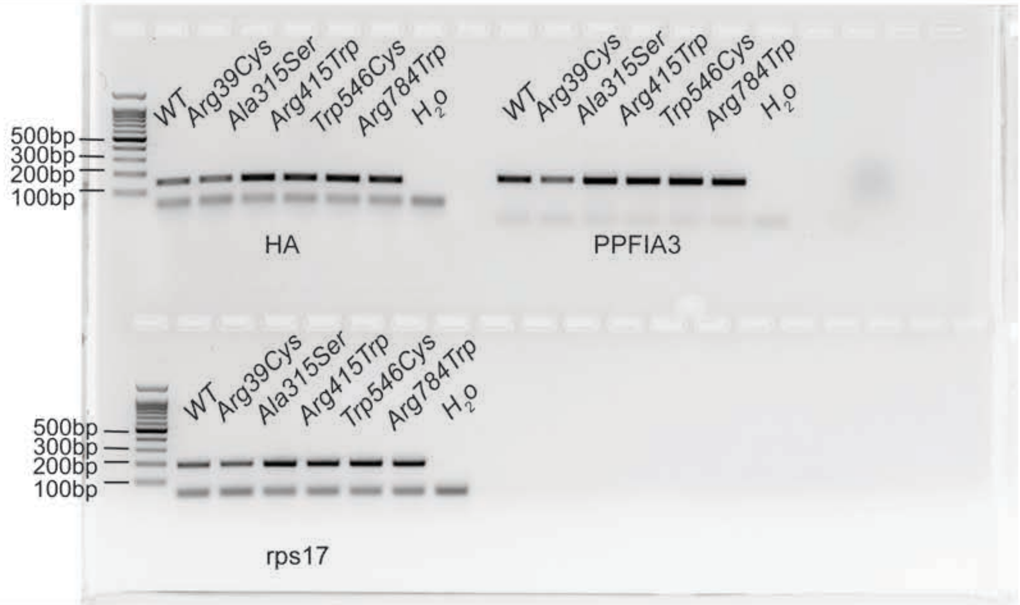


**B.** Sets 2 and 3 genomic DNA PCR raw image for quantifications in Figure S2

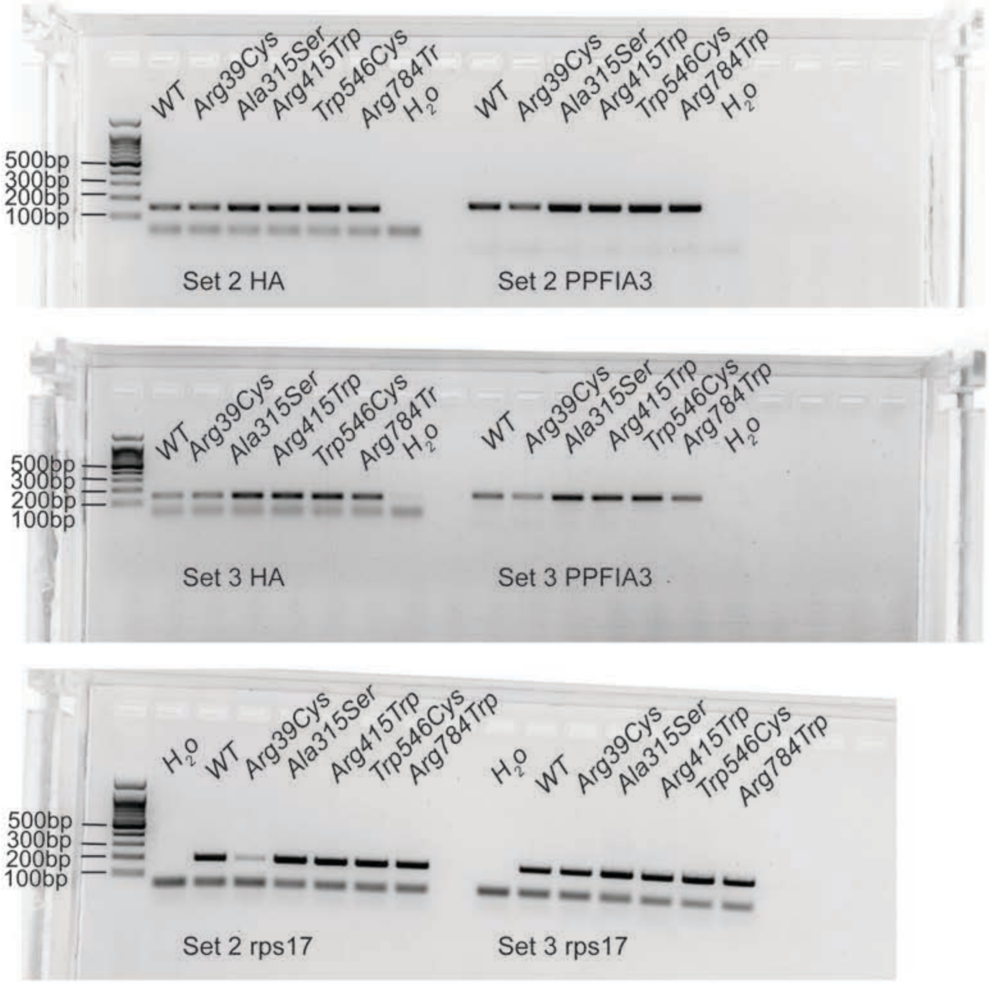

Figure S3

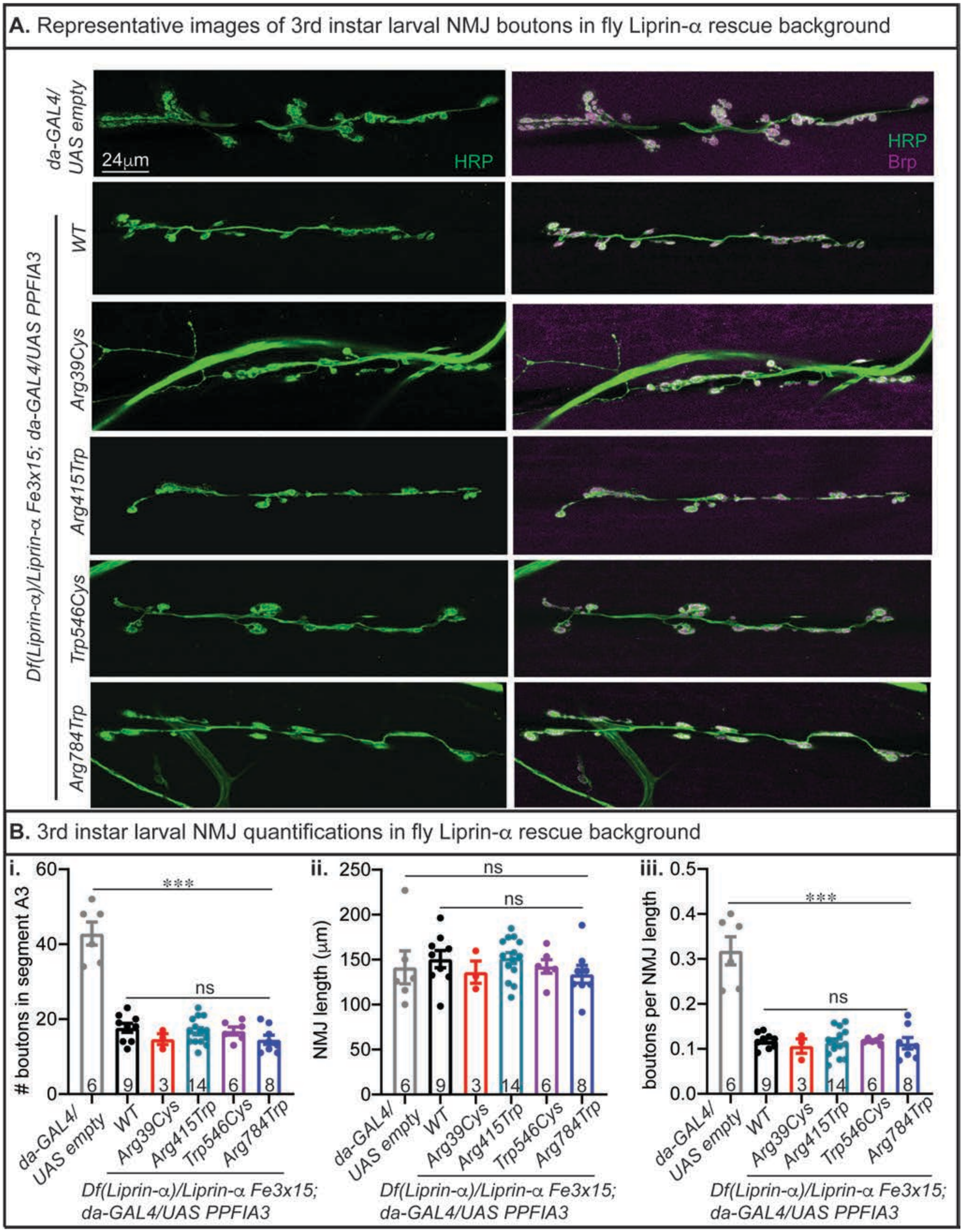

A. Representative images of 3rd instar larval NMJ boutons in fly Liprin-α rescue background
da-GAL4/ UAS empty
24μm
HRP
HRP
Brp
Df(Liprin-α)/Liprin-α Fe3x15; da-GAL4/UAS PPFIA3
WT
Arg39Cys
Arg415Trp
Trp546Cys
Arg784Trp
B. 3rd instar larval NMJ quantifications in fly Liprin-α rescue background
i.
### boutons in segment A3
***
ns
ii.
NMJ length (μm)
ns
ns
iii.
boutons per NMJ length
***
ns
6
9
3
14
6
8
da-GAL4/ UAS empty
WT
Arg39Cys
Arg415Trp
Trp546Cys
Arg784Trp
Df(Liprin-α)/Liprin-α Fe3x15; da-GAL4/UAS PPFIA3

**Table S1: Primer sequences for Q5 mutagenesis, sequencing, and genomic PCR**

| Primer name | Sequence | Purpose |
|---|---|---|
| PPFIA3 WT-NS-F | CTATTCCTGCGCGAACCCAGCTTTC | To remove stop codon from the WT cDNA |
| PPFIA3 WT-NS-R | GTCCGGACCGAAACGCCG | To remove stop codon from the WT cDNA |
| PPFIA3 p.Arg39Cys-NS-F | GCTCACGGAGTGCGAGC | Mutagenesis |
| PPFIA3 p.Arg39Cys -NS-R | ATCGTGACCATGAGGCGC | Mutagenesis |
| PPFIA3 p.Ala315Ser-NS-F | CTACCTGAGCTCCCAGCGGGA | Mutagenesis |
| PPFIA3 p.Ala315Ser-NS-R | CGCTTCTCCAGTGTTGTAATCC | Mutagenesis |
| PPFIA3 p.Arg415Trp-NS-F | TCAAGAGCTGCAGTGGGC | Mutagenesis |
| PPFIA3 p.Arg415Trp-NS-R | TTCTTCTCTTCCAGCTGGGCCTCC | Mutagenesis |
| PPFIA3 p.Trp546Cys-NS-F | GGGGCCGCTGTTCAGG | Mutagenesis |
| PPFIA3 p.Trp546Cys-NS-R | TCTTGCCTGCCCGAC | Mutagenesis |
| PPFIA3 p.Arg784Trp-NS-F | ACCCCCAGGCTGGGACAGCTC | Mutagenesis |
| PPFIA3 p.Arg784Trp-NS-R | CCCATTCGTCCCTTCTCTTTCTTG | Mutagenesis |
| PPFIA3 SeqP1 | TGTAAAACGACGGCCAGT | Sequencing |
| PPFIA3 SeqP2 | GGAGCGCGTGGCAGTGC | Sequencing |
| PPFIA3 SeqP3 | CGCTCAACAAGGCCGAGGAAC | Sequencing |
| PPFIA3 SeqP4 | GAGCAGCTGGAGGCCATCAAC | Sequencing |
| PPFIA3 SeqP5 | GGGCCTACCTTTTGCTGCC | Sequencing |
| PPFIA3 SeqP6 | CCTGAAGGAATTTGCCACG | Sequencing |
| PPFIA3 HA-F | TACCCATACGATGTTCCTGACTATG | PCR to check HA levels |
| PPFIA3 HA-R | AAGTGGCGCGACGCTTAGT | PCR to check HA levels |
| PPFIA3 cDNA-F | GAACAGCTGTCTAGGCGGCG | PCR to check PPFIA3 levels |
| PPFIA3 cDNA-R | CCGCAGCTGGCACACCT | PCR to check PPFIA3 levels |
| rps17-genomic-F | AAGCGCATCTGCGAGGAG | Endogenous control for PCR |
| rps17-genomic-R | CCTCCTCCTGCAACTTGATG | Endogenous control for PCR |

**Abbreviations:** wildtype (WT), no stop (NS), forward primer (F), reverse primer (R), sequencing primer (SeqP#)

**Table S2:** ***In silico*** **predictions for** ***PPFIA3*** **variants (individuals I:1 to I:14)**

| **Variant prediction** | **I:1** | **I:2** | **I:3** | **I:4** | **I:5** | **I:6** | **I:7** |
|---|---|---|---|---|---|---|---|
| Human reference genome | GRCh37 (hg19) | GRCh37 (hg19) | GRCh37 (hg19) | GRCh37 (hg19) | GRCh37 (hg19) | GRCh37 (hg19) | GRCh37 (hg19) |
| *PPFIA3* variant (cDNA) | NM_003660.4: c.115 C>T | NM_003660.4: c.239 A>C | NM_003660.4: c.240+1G>A | NM_003660.4: c.240+1G>A | NM_003660.4: c.943 G>T | NM_003660.4: c.1243 C>T | NM_003660.4: c.1285 C>T |
| PPFIA3 variant (protein) | NP_003651.1: p.(Arg39Cys) | NP_003651.1: p.(Gln80Pro) | n/a | n/a | NP_003651.1: p.(Ala315Ser) | NP_003651.1: p.(Arg415Trp) | NP_003651.1: p.(Arg429Trp) |
| CADD | 32 | 23.7 | 21 | 21 | 24.7 | 29.4 | 31 |
| GERP | 4.19 | 4.34 | 4.34 | 4.34 | 4.29 | 2.12 | 2.95 |
| M-CAP | Damaging | Damaging | n/a | n/a | Damaging | Tolerated | Damaging |
| PolyPhen2 HumDiv | Probably damaging | Benign | n/a | n/a | Benign | Probably Damaging | Probably damaging |
| PolyPhen2 HumVar | Probably damaging | Benign | n/a | n/a | Benign | Probably Damaging | Probably damaging |
| Phylop Vertebrate | 1.121 | 9.25 | n/a | n/a | 9.848 | 0.89 | 2.607 |
| SIFT | Damaging | Damaging | n/a | n/a | Damaging | Damaging | Damaging |

**Abbreviations:** combined annotation dependent depletion (CADD); genomic evolutionary rate profiling (GERP); Mendelian clinically applicable pathogenicity (M-CAP); polymorphism phenotyping v2 (PolyPhen2); sorting intolerant from tolerant (SIFT); not applicable (n/a)

| **Variant prediction** | **I:8** | **I:9** | **I:10** | **I:11** | **I:12** | **I:13** | **I:14** |
|---|---|---|---|---|---|---|---|
| Human reference genome | GRCh37 (hg19) | GRCh37 (hg19) | GRCh37 (hg19) | GRCh37 (hg19) | GRCh37 (hg19) | GRCh37 (hg19) | GRCh37 (hg19) |
| *PPFIA3* variant (cDNA) | NM_003660.4: c.1492 C>T | NM_003660.4: c.1638 G>T | NM_003660.4: c.2350 C>T | NM_003660.4: c.2609 T>A | NM_003660.4: c.2609 T>A | NM_003660.4: c.2717 C>T | NM_003660.4: c.3307del |
| *PPFIA3* variant (protein) | NP_003651.1: p.(Arg498Trp) | NP_003651.1: p.(Trp546Cys) | NP_003651.1: p.(Arg784Trp) | NP_003651.1: p.(Ile870Asn) | NP_003651.1: p.(Ile870Asn) | NP_003651.1: p.(Ser906Leu) | NP_003651.1: p.(Glu1103Asnfs*8) |
| CADD | 25.3 | 27.5 | 24.4 | 35 | 35 | 28.8 | n/a |
| GERP | 1.41 | 3.87 | 3.63 | 4.45 | 4.45 | 3.31 | 4.29 |
| M-CAP | Damaging | Damaging | Damaging | Damaging | Damaging | Probably damaging | n/a |
| PolyPhen2 HumDiv | Probably damaging | Probably damaging | Probably damaging | Probably damaging | Probably damaging | Tolerated | n/a |
| PolyPhen2 HumVar | Probably damaging | Probably damaging | Benign | Probably damaging | Probably damaging | Probably damaging | n/a |
| Phylop Vertebrate | 2.132 | 4.461 | 1.309 | 7.965 | 7.965 | 5.913 | n/a |
| SIFT | Damaging | Tolerated | Damaging | Damaging | Damaging | Damaging | n/a |

**Abbreviations:** combined annotation dependent depletion (CADD); genomic evolutionary rate profiling (GERP); Mendelian clinically applicable pathogenicity (M-CAP); polymorphism phenotyping v2 (PolyPhen2); sorting intolerant from tolerant (SIFT); not applicable (n/a)

**Table S6: MRI brain findings in individuals I:1 - I:14 with *PPFIA3* variants**

| **MRI Brain** | **I:1** | **I:2** | **I:3** | **I:4** | **I:5** | **I:6** | **I:7** |
|---|---|---|---|---|---|---|---|
| *PPFIA3* variant (cDNA) | NM_003660.4: c.115 C>T | NM_003660.4: c.239 A>C | NM_003660.4: c.240+1 G>A | NM_003660.4: c.240+1 G>A | NM_003660.4: c.943 G>T | NM_003660.4: c.1243 C>T | NM_003660.4: c.1285 C>T |
| PPFIA3 variant (protein) | NP_003651.1: p.(Arg39Cys) | NP_003651.1: p.(Gln80Pro) | n/a | n/a | NP_003651.1: p.(Ala315Ser) | NP_003651.1: p.(Arg415Trp) | NP_003651.1: p.(Arg429Trp) |
| Cerebral volume loss | no | no | no | n/a | n/a | no | n/a |
| T2 hyperintensity | no | no | no | n/a | n/a | no | n/a |
| Contrast enhancement | no | no | no | n/a | n/a | no | n/a |
| Diffusion restriction | no | no | no | n/a | n/a | no | n/a |
| Hypomyelination | no | no | no | n/a | n/a | no | n/a |
| Thinning of the corpus callosum | no | no | no | n/a | n/a | no | n/a |
| Vermis volume loss | no | no | no | n/a | n/a | no | n/a |
| Additional MRI features | Normal brain MRI with no seizure locus identified | normal study | normal study | n/a | n/a | Flattening of the posterior globes at the level of the insertion of the optic nerves | n/a |

**Abbreviations:** magnetic resonance imaging (MRI), no information available (n/a)

| MRI Brain | I:8 | I:9 | I:10 | I:11 | I:12 | I:13 | I:14 |
|---|---|---|---|---|---|---|---|
| *PPFIA3* variant (cDNA) | NM_003660.4: c.1492 C>T | NM_003660.4: c.1638 G>T | NM_003660.4: c.2350 C>T | NM_003660.4: c.2609 T>A | NM_003660.4: c.2609 T>A | NM_003660.4: c.2717 C>T | NM_003660.4: c.3307del |
| PPFIA3 variant (protein) | NP_003651.1: p.(Arg498Trp) | NP_003651.1: p.(Trp546Cys) | NP_003651.1: p.(Arg784Trp) | NP_003651.1: p.(Ile870Asn) | NP_003651.1: p.(Ile870Asn) | NP_003651.1: p.(Ser906Leu) | NP_003651.1: p.(Glu1103Asnfs*8) |
| Cerebral volume loss | n/a | no | no | n/a | no | no | no |
| T2 hyperintensity | n/a | no | no | n/a | no | no | no |
| Contrast enhancement | n/a | no | no | n/a | no | no | no |
| Diffusion restriction | n/a | no | no | n/a | no | no | no |
| Hypomyelination | n/a | no | no | n/a | no | no | no |
| Thinning of the corpus callosum | n/a | no | no | n/a | no | no | no |
| Vermis volume loss | n/a | no | no | n/a | no | no | no |
| Additional MRI features | n/a | normal study | normal study | n/a | Significant right posterior plagiocephaly; mild periventricular signal abnormality with associated mild white matter volume loss, suggesting chronic sequelae of mild periventricular leukomalacia. | normal study | normal study |

**Abbreviations:** magnetic resonance imaging (MRI), no information available (n/a)

**Table S7: Statistical summary of data for *PPFIA3* variant studies**

| Figure | Data Type | Measurement | Statistical Test | Comparison | Statistics | DFn | DFd | *p* |
|---|---|---|---|---|---|---|---|---|
| **Figure 4, A** | behavior | climbing (elav-Gal4 > UAS lines) | one-way ANOVA | both genders, all genotypes | F =9.208 | 6 | 339 | <0.0001 |
| | | | Tukey's post-hoc analysis | empty vs. WT | q = 1.069 | n/a | 339 | 0.9887 |
| | | | Tukey's post-hoc analysis | empty vs. R39C | q = 8.902 | n/a | 339 | <0.0001 |
| | | | Tukey's post-hoc analysis | empty vs. A315S | q = 3.06 | n/a | 339 | 0.3183 |
| | | | Tukey's post-hoc analysis | empty vs. R415W | q = 6.04 | n/a | 339 | 0.0005 |
| | | | Tukey's post-hoc analysis | empty vs. W546C | q = 4.242 | n/a | 339 | 0.0455 |
| | | | Tukey's post-hoc analysis | empty vs. R784W | q = 4.896 | n/a | 339 | 0.0107 |
| | | | Tukey's post-hoc analysis | WT vs. R39C | q = 7.973 | n/a | 339 | <0.0001 |
| | | | Tukey's post-hoc analysis | WT vs. A315S | q = 2.038 | n/a | 339 | 0.7791 |
| | | | Tukey's post-hoc analysis | WT vs. R415W | q = 5.068 | n/a | 339 | 0.0071 |
| | | | Tukey's post-hoc analysis | WT vs. W546C | q = 3.285 | n/a | 339 | 0.2361 |
| | | | Tukey's post-hoc analysis | WT vs. R784W | q = 3.91 | n/a | 339 | 0.0861 |
| | | | Tukey's post-hoc analysis | R39C vs. A315S | q = 5.862 | n/a | 339 | 0.0008 |
| | | | Tukey's post-hoc analysis | R39C vs. R415W | q = 2.852 | n/a | 339 | 0.4059 |
| | | | Tukey's post-hoc analysis | R39C vs. W546C | q = 4.268 | n/a | 339 | 0.0432 |
| | | | Tukey's post-hoc analysis | R39C vs. R784W | q = 3.973 | n/a | 339 | 0.0766 |
| | | | Tukey's post-hoc analysis | A315S vs. R415W | q = 2.997 | n/a | 339 | 0.3437 |
| | | | Tukey's post-hoc analysis | A315S vs. W546C | q = 1.33 | n/a | 339 | 0.9657 |
| | | | Tukey's post-hoc analysis | A315S vs. R784W | q = 1.859 | n/a | 339 | 0.8449 |
| | | | Tukey's post-hoc analysis | R415W vs. W546C | q = 1.534 | n/a | 339 | 0.9324 |
| | | | Tukey's post-hoc analysis | R415W vs. R784W | q = 1.127 | n/a | 339 | 0.9852 |
| | | | Tukey's post-hoc analysis | W546C vs. R784W | q = 0.4511 | n/a | 339 | >0.9999 |

| | | | | | | | | |
|---|---|---|---|---|---|---|---|---|
| **Figure 4, B** | behavior | bang sensitivity (elav-Gal4 > UAS lines) | one-way ANOVA | both genders, all genotypes | F = 25.37 | 6 | 337 | <0.0001 |
| | | | Tukey's post-hoc analysis | empty vs. WT | q = 1.705 | n/a | 337 | 0.8919 |
| | | | Tukey's post-hoc analysis | empty vs. R39C | q = 10.84 | n/a | 337 | <0.0001 |
| | | | Tukey's post-hoc analysis | empty vs. A315S | q = 11.61 | n/a | 337 | <0.0001 |
| | | | Tukey's post-hoc analysis | empty vs. R415W | q = 12.17 | n/a | 337 | <0.0001 |
| | | | Tukey's post-hoc analysis | empty vs. W546C | q = 3.572 | n/a | 337 | 0.1534 |
| | | | Tukey's post-hoc analysis | empty vs. R784W | q = 4.761 | n/a | 337 | 0.0147 |
| | | | Tukey's post-hoc analysis | WT vs. R39C | q = 9.191 | n/a | 337 | <0.0001 |
| | | | Tukey's post-hoc analysis | WT vs. A315S | q = 9.983 | n/a | 337 | <0.0001 |
| | | | Tukey's post-hoc analysis | WT vs. R415W | q = 10.53 | n/a | 337 | <0.0001 |
| | | | Tukey's post-hoc analysis | WT vs. W546C | q = 1.979 | n/a | 337 | 0.802 |
| | | | Tukey's post-hoc analysis | WT vs. R784W | q = 3.071 | n/a | 337 | 0.3137 |
| | | | Tukey's post-hoc analysis | R39C vs. A315S | q = 0.8794 | n/a | 337 | 0.9961 |
| | | | Tukey's post-hoc analysis | R39C vs. R415W | q = 1.325 | n/a | 337 | 0.9663 |
| | | | Tukey's post-hoc analysis | R39C vs. W546C | q = 6.679 | n/a | 337 | <0.0001 |
| | | | Tukey's post-hoc analysis | R39C vs. R784W | q = 6.15 | n/a | 337 | 0.0004 |
| | | | Tukey's post-hoc analysis | A315S vs. R415W | q = 0.4322 | n/a | 337 | >0.9999 |
| | | | Tukey's post-hoc analysis | A315S vs. W546C | q = 7.453 | n/a | 337 | <0.0001 |
| | | | Tukey's post-hoc analysis | A315S vs. R784W | q = 6.974 | n/a | 337 | <0.0001 |
| | | | Tukey's post-hoc analysis | R415W vs. W546C | q = 7.937 | n/a | 337 | <0.0001 |
| | | | Tukey's post-hoc analysis | R415W vs. R784W | q = 7.488 | n/a | 337 | <0.0001 |
| | | | Tukey's post-hoc analysis | W546C vs. R784W | q = 0.905 | n/a | 337 | 0.9954 |
| **Figure 4, C, iii** | NMJ morphology | NMJ boutons (elav-Gal4 > UAS lines) | one-way ANOVA | both genders, all genotypes | F = 13.54 | 5 | 127 | <0.0001 |
| | | | Tukey's post-hoc analysis | empty vs. WT | q = 0.7621 | n/a | 127 | 0.9944 |

| | | | | | | | |
|---|---|---|---|---|---|---|---|
| | | | Tukey's post-hoc analysis | empty vs. R39C | q = 6.397 | n/a | 127 | 0.0002 |
| | | | Tukey's post-hoc analysis | empty vs. R415W | q = 5.613 | n/a | 127 | 0.0016 |
| | | | Tukey's post-hoc analysis | empty vs. R784W | q = 1.612 | n/a | 127 | 0.8637 |
| | | | Tukey's post-hoc analysis | empty vs. W546C | q = 2.873 | n/a | 127 | 0.3306 |
| | | | Tukey's post-hoc analysis | WT vs. R39C | q = 6.23 | n/a | 127 | 0.0003 |
| | | | Tukey's post-hoc analysis | WT vs. R415W | q = 5.351 | n/a | 127 | 0.0032 |
| | | | Tukey's post-hoc analysis | WT vs. R784W | q = 2.39 | n/a | 127 | 0.5409 |
| | | | Tukey's post-hoc analysis | WT vs. W546C | q = 3.847 | n/a | 127 | 0.0783 |
| | | | Tukey's post-hoc analysis | R39C vs. R415W | q = 0.2249 | n/a | 127 | >0.9999 |
| | | | Tukey's post-hoc analysis | R39C vs. R784W | q = 7.297 | n/a | 127 | <0.0001 |
| | | | Tukey's post-hoc analysis | R39C vs. W546C | q = 9.066 | n/a | 127 | <0.0001 |
| | | | Tukey's post-hoc analysis | R415W vs. R784W | q = 6.579 | n/a | 127 | 0.0001 |
| | | | Tukey's post-hoc analysis | R415W vs. W546C | q = 8.105 | n/a | 127 | <0.0001 |
| | | | Tukey's post-hoc analysis | R784W vs. W546C | q = 1.005 | n/a | 127 | 0.9804 |
| **Figure 4, C, iv** | NMJ morphology | NMJ length (elav-Gal4 > UAS lines) | one-way ANOVA | both genders, all genotypes | F = 2.314 | 5 | 127 | 0.0475 |
| | | | Tukey's post-hoc analysis | empty vs. WT | q = 0.2255 | n/a | 127 | >0.9999 |
| | | | Tukey's post-hoc analysis | empty vs. R39C | q = 2.045 | n/a | 127 | 0.6988 |
| | | | Tukey's post-hoc analysis | empty vs. R415W | q = 2.84 | n/a | 127 | 0.3433 |
| | | | Tukey's post-hoc analysis | empty vs. W546C | q = 1.044 | n/a | 127 | 0.9767 |
| | | | Tukey's post-hoc analysis | empty vs. R784W | q = 0.6501 | n/a | 127 | 0.9974 |
| | | | Tukey's post-hoc analysis | WT vs. R39C | q = 2.453 | n/a | 127 | 0.5119 |
| | | | Tukey's post-hoc analysis | WT vs. R415W | q = 3.26 | n/a | 127 | 0.1998 |
| | | | Tukey's post-hoc analysis | WT vs. W546C | q = 0.9105 | n/a | 127 | 0.9874 |
| | | | Tukey's post-hoc analysis | WT vs. R784W | q = 0.4962 | n/a | 127 | 0.9993 |

| | | | | | | | | |
|---|---|---|---|---|---|---|---|---|
| | | | Tukey's post-hoc analysis | R39C vs. R415W | q = 0.953 | n/a | 127 | 0.9845 |
| | | | Tukey's post-hoc analysis | R39C vs. W546C | q = 3.025 | n/a | 127 | 0.2741 |
| | | | Tukey's post-hoc analysis | R39C vs. R784W | q = 2.469 | n/a | 127 | 0.5046 |
| | | | Tukey's post-hoc analysis | R415W vs. W546C | q = 3.734 | n/a | 127 | 0.0952 |
| | | | Tukey's post-hoc analysis | R415W vs. R784W | q = 3.174 | n/a | 127 | 0.2252 |
| | | | Tukey's post-hoc analysis | W546C vs. R784W | q = 0.303 | n/a | 127 | >0.9999 |
| **Figure 5, A, ii** | liprin-a LOF lethality rescue | larval stage rescue (da-Gal4 > UAS lines) | one-way ANOVA | both genders, all genotypes | F = 7.922 | 5 | 12 | 0.0017 |
| | | | Tukey's post-hoc analysis | WT vs. R39C | q = 6.778 | n/a | 12 | 0.0045 |
| | | | Tukey's post-hoc analysis | WT vs. R415W | q = 4.763 | n/a | 12 | 0.0492 |
| | | | Tukey's post-hoc analysis | WT vs. R784W | q = 3.485 | n/a | 12 | 0.2094 |
| | | | Tukey's post-hoc analysis | WT vs. W546C | q = 3.345 | n/a | 12 | 0.2419 |
| | | | Tukey's post-hoc analysis | WT vs. empty | q = 7.938 | n/a | 12 | 0.0012 |
| | | | Tukey's post-hoc analysis | R39C vs. R415W | q = 2.014 | n/a | 12 | 0.7133 |
| | | | Tukey's post-hoc analysis | R39C vs. R784W | q = 3.293 | n/a | 12 | 0.2548 |
| | | | Tukey's post-hoc analysis | R39C vs. W546C | q = 3.433 | n/a | 12 | 0.2209 |
| | | | Tukey's post-hoc analysis | R39C vs. empty | q = 1.16 | n/a | 12 | 0.958 |
| | | | Tukey's post-hoc analysis | R415W vs. R784W | q = 1.279 | n/a | 12 | 0.9381 |
| | | | Tukey's post-hoc analysis | R415W vs. W546C | q = 1.419 | n/a | 12 | 0.9082 |
| | | | Tukey's post-hoc analysis | R415W vs. empty | q = 3.174 | n/a | 12 | 0.2867 |
| | | | Tukey's post-hoc analysis | R784W vs. W546C | q = 0.1398 | n/a | 12 | >0.9999 |
| | | | Tukey's post-hoc analysis | R784W vs. empty | q = 4.453 | n/a | 12 | 0.0709 |
| | | | Tukey's post-hoc analysis | \W546C vs. empty | q = 4.593 | n/a | 12 | 0.0602 |
| **Figure 5, B, ii** | liprin-a LOF lethality rescue | adult stage rescue (da-Gal4 > UAS lines) | one-way ANOVA | both genders, all genotypes | F = 51.50 | 5 | 12 | <0.0001 |
| | | | Tukey's post-hoc analysis | WT vs. R39C | q = 15.88 | n/a | 12 | <0.0001 |

| | | | | | | | | |
|---|---|---|---|---|---|---|---|---|
| | | | Tukey's post-hoc analysis | WT vs. R415W | q = 5.287 | n/a | 12 | 0.0264 |
| | | | Tukey's post-hoc analysis | WT vs. R784W | q = 2.266 | n/a | 12 | 0.612 |
| | | | Tukey's post-hoc analysis | WT vs. W546C | q = 0.7906 | n/a | 12 | 0.992 |
| | | | Tukey's post-hoc analysis | WT vs. empty | q = 15.18 | n/a | 12 | <0.0001 |
| | | | Tukey's post-hoc analysis | R39C vs. R415W | q = 10.6 | n/a | 12 | <0.0001 |
| | | | Tukey's post-hoc analysis | R39C vs. R784W | q = 13.62 | n/a | 12 | <0.0001 |
| | | | Tukey's post-hoc analysis | R39C vs. W546C | q = 15.09 | n/a | 12 | <0.0001 |
| | | | Tukey's post-hoc analysis | R39C vs. empty | q = 0.7074 | n/a | 12 | 0.9952 |
| | | | Tukey's post-hoc analysis | R415W vs. R784W | q = 3.021 | n/a | 12 | 0.332 |
| | | | Tukey's post-hoc analysis | R415W vs. W546C | q = 4.497 | n/a | 12 | 0.0674 |
| | | | Tukey's post-hoc analysis | R415W vs. empty | q = 9.888 | n/a | 12 | 0.0002 |
| | | | Tukey's post-hoc analysis | R784W vs. W546C | q = 1.476 | n/a | 12 | 0.894 |
| | | | Tukey's post-hoc analysis | R784W vs. empty | q = 12.91 | n/a | 12 | <0.0001 |
| | | | Tukey's post-hoc analysis | \W546C vs. empty | q = 14.38 | n/a | 12 | <0.0001 |
| **Figure 5, B, iii** | liprin-a LOF lethality rescue | rescue adult viability (da-Gal4 > UAS lines) | one-way ANOVA | both genders, all genotypes | F = 12.67 | 4 | 8 | 0.0015 |
| | | | Tukey's post-hoc analysis | WT vs. R415W | q = 5.694 | n/a | 8 | 0.0232 |
| | | | Tukey's post-hoc analysis | WT vs. R784W | q = 6.707 | n/a | 8 | 0.0093 |
| | | | Tukey's post-hoc analysis | WT vs. W546C | q = 1.28 | n/a | 8 | 0.8873 |
| | | | Tukey's post-hoc analysis | WT vs. empty | q = 7.893 | n/a | 8 | 0.0034 |
| | | | Tukey's post-hoc analysis | R415W vs. R784W | q = 1.013 | n/a | 8 | 0.9469 |
| | | | Tukey's post-hoc analysis | R415W vs. W546C | q = 4.414 | n/a | 8 | 0.0789 |
| | | | Tukey's post-hoc analysis | R415W vs. empty | q = 3.867 | n/a | 8 | 0.1337 |
| | | | Tukey's post-hoc analysis | R784W vs. W546C | q = 5.427 | n/a | 8 | 0.0299 |
| | | | Tukey's post-hoc analysis | R784W vs. empty | q = 3.151 | n/a | 8 | 0.2605 |

| | | | | | | | | |
|---|---|---|---|---|---|---|---|---|
| | | | Tukey's post-hoc analysis | \W546C vs. empty | q = 6.988 | n/a | 8 | 0.0073 |
| **Figure S1, C, ii** | genomic DNA PCR | relative HA genomic DNA levels | one-way ANOVA | both genders, all genotypes | F = 0.7202 | 5 | 12 | 0.6208 |
| | | | Tukey's post-hoc analysis | WT vs. R39C | q = 0.4298 | n/a | 12 | 0.9996 |
| | | | Tukey's post-hoc analysis | WT vs. R315S | q = 1.075 | n/a | 12 | 0.9693 |
| | | | Tukey's post-hoc analysis | WT vs. R415W | q = 1.92 | n/a | 12 | 0.7498 |
| | | | Tukey's post-hoc analysis | WT vs. W546C | q = 1.176 | n/a | 12 | 0.9556 |
| | | | Tukey's post-hoc analysis | WT vs. R784W | q = 2.228 | n/a | 12 | 0.6276 |
| | | | Tukey's post-hoc analysis | R39C vs. R315S | q = 0.6453 | n/a | 12 | 0.9969 |
| | | | Tukey's post-hoc analysis | R39C vs. R415W | q = 1.49 | n/a | 12 | 0.8904 |
| | | | Tukey's post-hoc analysis | R39C vs. W546C | q = 0.7462 | n/a | 12 | 0.9939 |
| | | | Tukey's post-hoc analysis | R39C vs. R784W | q = 1.798 | n/a | 12 | 0.7943 |
| | | | Tukey's post-hoc analysis | R315S vs. R415W | q = 0.8445 | n/a | 12 | 0.9892 |
| | | | Tukey's post-hoc analysis | R315S vs. W546C | q = 0.1009 | n/a | 12 | >0.9999 |
| | | | Tukey's post-hoc analysis | R315S vs. R784W | q = 1.153 | n/a | 12 | 0.9591 |
| | | | Tukey's post-hoc analysis | R415W vs. W546C | q = 0.7436 | n/a | 12 | 0.994 |
| | | | Tukey's post-hoc analysis | R415W vs. R784W | q = 0.3083 | n/a | 12 | >0.9999 |
| | | | Tukey's post-hoc analysis | W546C vs. R784W | q = 1.052 | n/a | 12 | 0.972 |
| **Figure S1, C, iii** | genomic DNA PCR | relative PPFIA3 genomic DNA levels | one-way ANOVA | both genders, all genotypes | F = 1.305 | 5 | 12 | 0.3252 |
| | | | Tukey's post-hoc analysis | WT vs. R39C | q = 3.481 | n/a | 12 | 0.2101 |
| | | | Tukey's post-hoc analysis | WT vs. R315S | q = 1.735 | n/a | 12 | 0.8164 |
| | | | Tukey's post-hoc analysis | WT vs. R415W | q = 1.288 | n/a | 12 | 0.9364 |
| | | | Tukey's post-hoc analysis | WT vs. W546C | q = 2.104 | n/a | 12 | 0.6779 |
| | | | Tukey's post-hoc analysis | WT vs. R784W | q = 1.326 | n/a | 12 | 0.9287 |
| | | | Tukey's post-hoc analysis | R39C vs. R315S | q = 1.747 | n/a | 12 | 0.8122 |

| | | | | | | | | |
|---|---|---|---|---|---|---|---|---|
| | | | Tukey's post-hoc analysis | R39C vs. R415W | q = 2.193 | n/a | 12 | 0.6416 |
| | | | Tukey's post-hoc analysis | R39C vs. W546C | q = 1.378 | n/a | 12 | 0.9177 |
| | | | Tukey's post-hoc analysis | R39C vs. R784W | q = 2.155 | n/a | 12 | 0.6573 |
| | | | Tukey's post-hoc analysis | R315S vs. R415W | q = 0.4467 | n/a | 12 | 0.9995 |
| | | | Tukey's post-hoc analysis | R315S vs. W546C | q = 0.369 | n/a | 12 | 0.9998 |
| | | | Tukey's post-hoc analysis | R315S vs. R784W | q = 0.4081 | n/a | 12 | 0.9997 |
| | | | Tukey's post-hoc analysis | R415W vs. W546C | q = 0.8157 | n/a | 12 | 0.9908 |
| | | | Tukey's post-hoc analysis | R415W vs. R784W | q = 0.03865 | n/a | 12 | >0.9999 |
| | | | Tukey's post-hoc analysis | W546C vs. R784W | q = 0.777 | n/a | 12 | 0.9926 |
| **Figure S3, B, i** | NMJ morphology | rescue NMJ boutons (da-GAL4 > UAS lines) | one-way ANOVA | both genders, all genotypes | F = 31.45 | 5 | 40 | P<0.0001 |
| | | | Tukey's post-hoc analysis | da GAL4/empty vs. WT | q = 14.46 | n/a | 40 | <0.0001 |
| | | | Tukey's post-hoc analysis | da GAL4/empty vs. R39C | q = 11.41 | n/a | 40 | <0.0001 |
| | | | Tukey's post-hoc analysis | da GAL4/empty vs. R415W | q = 15.95 | n/a | 40 | <0.0001 |
| | | | Tukey's post-hoc analysis | da GAL4/empty vs. W546C | q = 13.21 | n/a | 40 | <0.0001 |
| | | | Tukey's post-hoc analysis | da GAL4/empty vs. R784W | q = 14.52 | n/a | 40 | <0.0001 |
| | | | Tukey's post-hoc analysis | WT vs. R39C | q = 0.678 | n/a | 40 | 0.9966 |
| | | | Tukey's post-hoc analysis | WT vs. R415W | q = 0.3831 | n/a | 40 | 0.9998 |
| | | | Tukey's post-hoc analysis | WT vs. W546C | q = 0.01587 | n/a | 40 | >0.9999 |
| | | | Tukey's post-hoc analysis | WT vs. R784W | q = 0.4554 | n/a | 40 | 0.9995 |
| | | | Tukey's post-hoc analysis | R39C vs. R415W | q = 0.4531 | n/a | 40 | 0.9995 |
| | | | Tukey's post-hoc analysis | R39C vs. W546C | q = 0.6274 | n/a | 40 | 0.9977 |
| | | | Tukey's post-hoc analysis | R39C vs. R784W | q = 0.3407 | n/a | 40 | 0.9999 |
| | | | Tukey's post-hoc analysis | R415W vs. W546C | q = 0.3183 | n/a | 40 | >0.9999 |
| | | | Tukey's post-hoc analysis | R415W vs. R784W | q = 0.13 | n/a | 40 | >0.9999 |

| | | | | | | | | |
|---|---|---|---|---|---|---|---|---|
| | | | Tukey's post-hoc analysis | W546C vs. R784W | q = 0.3943 | n/a | 40 | 0.9998 |
| **Figure S3, B, ii** | NMJ morphology | rescue NMJ length (da-GAL4 > UAS lines) | one-way ANOVA | both genders, all genotypes | F = 0.5746 | 5 | 40 | P=0.7190 |
| | | | Tukey's post-hoc analysis | da GAL4/empty vs. WT | q = 0.8756 | n/a | 40 | 0.989 |
| | | | Tukey's post-hoc analysis | da GAL4/empty vs. R39C | q = 0.3709 | n/a | 40 | 0.9998 |
| | | | Tukey's post-hoc analysis | da GAL4/empty vs. R415W | q = 1.067 | n/a | 40 | 0.9734 |
| | | | Tukey's post-hoc analysis | da GAL4/empty vs. W546C | q = 0.0938 | n/a | 40 | >0.9999 |
| | | | Tukey's post-hoc analysis | da GAL4/empty vs. R784W | q = 0.7085 | n/a | 40 | 0.9959 |
| | | | Tukey's post-hoc analysis | WT vs. R39C | q = 1.086 | n/a | 40 | 0.9714 |
| | | | Tukey's post-hoc analysis | WT vs. R415W | q = 0.1389 | n/a | 40 | >0.9999 |
| | | | Tukey's post-hoc analysis | WT vs. W546C | q = 0.7729 | n/a | 40 | 0.9938 |
| | | | Tukey's post-hoc analysis | WT vs. R784W | q = 1.737 | n/a | 40 | 0.8204 |
| | | | Tukey's post-hoc analysis | R39C vs. R415W | q = 1.231 | n/a | 40 | 0.9514 |
| | | | Tukey's post-hoc analysis | R39C vs. W546C | q = 0.4475 | n/a | 40 | 0.9995 |
| | | | Tukey's post-hoc analysis | R39C vs. R784W | q = 0.1778 | n/a | 40 | >0.9999 |
| | | | Tukey's post-hoc analysis | R415W vs. W546C | q = 0.9565 | n/a | 40 | 0.9836 |
| | | | Tukey's post-hoc analysis | R415W vs. R784W | q = 2.039 | n/a | 40 | 0.702 |
| | | | Tukey's post-hoc analysis | W546C vs. R784W | q = 0.8088 | n/a | 40 | 0.9923 |
| **Figure S3, B, iii** | NMJ morphology | rescue NMJ boutons/length (da-GAL4 > UAS lines) | one-way ANOVA | both genders, all genotypes | F = 31.45 | 5 | 40 | P<0.0001 |
| | | | Tukey's post-hoc analysis | da GAL4/empty vs. WT | q = 14.46 | n/a | 40 | <0.0001 |
| | | | Tukey's post-hoc analysis | da GAL4/empty vs. R39C | q = 11.41 | n/a | 40 | <0.0001 |
| | | | Tukey's post-hoc analysis | da GAL4/empty vs. R415W | q = 15.95 | n/a | 40 | <0.0001 |
| | | | Tukey's post-hoc analysis | da GAL4/empty vs. W546C | q = 13.21 | n/a | 40 | <0.0001 |
| | | | Tukey's post-hoc analysis | da GAL4/empty vs. R784W | q = 14.52 | n/a | 40 | <0.0001 |
| | | | Tukey's post-hoc analysis | WT vs. R39C | q = 0.678 | n/a | 40 | 0.9966 |

| | | | | | | | |
|---|---|---|---|---|---|---|---|
| | | Tukey's post-hoc analysis | WT vs. R415W | q = 0.3831 | n/a | 40 | 0.9998 |
| | | Tukey's post-hoc analysis | WT vs. W546C | q = 0.01587 | n/a | 40 | >0.9999 |
| | | Tukey's post-hoc analysis | WT vs. R784W | q = 0.4554 | n/a | 40 | 0.9995 |
| | | Tukey's post-hoc analysis | R39C vs. R415W | q = 0.4531 | n/a | 40 | 0.9995 |
| | | Tukey's post-hoc analysis | R39C vs. W546C | q = 0.6274 | n/a | 40 | 0.9977 |
| | | Tukey's post-hoc analysis | R39C vs. R784W | q = 0.3407 | n/a | 40 | 0.9999 |
| | | Tukey's post-hoc analysis | R415W vs. W546C | q = 0.3183 | n/a | 40 | >0.9999 |
| | | Tukey's post-hoc analysis | R415W vs. R784W | q = 0.13 | n/a | 40 | >0.9999 |
| | | Tukey's post-hoc analysis | W546C vs. R784W | q = 0.3943 | n/a | 40 | 0.9998 |
